# CT radiomics to predict checkpoint inhibitors treatment outcomes in patients with advanced cutaneous melanoma

**DOI:** 10.1101/2022.12.19.22283574

**Authors:** L.S. ter Maat, I.A.J. van Duin, S.G. Elias, T. Leiner, J.J.C. Verhoeff, E.R.A.N. Arntz, M.F. Troenokarso, W.A.M. Blokx, I. Isgum, G.A. de Wit, F.W.P.J. van den Berkmortel, M.J. Boers-Sonderen, M.F. Boomsma, A.J.M. van den Eertwegh, J.W.B. de Groot, D. Piersma, G. Vreugdenhil, H.M Westgeest, E. Kapiteijn, P.J. van Diest, J.P.W. Pluim, P.A. de Jong, K.P.M. Suijkerbuijk, M. Veta

## Abstract

**Introduction:** Predicting checkpoint inhibitors treatment outcomes in melanoma is a relevant task, due to the unpredictable and potentially fatal toxicity and high costs for society. However, accurate biomarkers for treatment outcomes are lacking. Radiomics are a technique to quantitatively capture tumor characteristics on readily available computed tomography (CT) imaging. The purpose of this study was to investigate the added value of radiomics for predicting durable clinical benefit from checkpoint inhibitors in melanoma in a large, multicenter cohort.

**Methods:** Patients who received first-line anti-PD1 ± anti-CTLA4 treatment for advanced cutaneous melanoma were retrospectively identified from nine participating hospitals. For every patient, up to five representative lesions were segmented on baseline CT and radiomics features were extracted. A machine learning pipeline was trained on the radiomics features to predict durable clinical benefit, defined as stable disease for more than six months or response per RECIST 1.1 criteria. This approach was evaluated using a leave-one-center-out cross validation and compared to a model based on previously discovered clinical predictors. Lastly, a combination model was built on the radiomics and clinical model.

**Results:** A total of 620 patients were included, of which 59.2% experienced durable clinical benefit. The radiomics model achieved an area under the receiver operator characteristic curve (AUROC) of 0.607 [95%CI 0.562-0.652], lower than that of the clinical model (AUROC=0.646 [95%CI 0.600-0.692]). The combination model yielded no improvement over the clinical model in terms of discrimination (AUROC=0.636 [95%CI 0.592-0.680]) or calibration. The output of the radiomics model was significantly correlated with three out of five input variables of the clinical model (p < 0.001).

**Discussion:** The radiomics model achieved a moderate predictive value of durable clinical benefit, which was statistically significant. However, a radiomics approach was unable to add value to a simpler clinical model, most likely due to the overlap in predictive information learned by both models. Future research should focus on the application of deep learning, spectral CT derived radiomics and a multimodal approach for accurately predicting benefit to checkpoint inhibitor treatment in advanced melanoma.

## Introduction

### Survival of patients with advanced melanoma has improved dramatically after the introduction of immunotherapy

The survival of patients with unresectable stage IIIC and stage IV melanoma has historically been very poor with a 1-year overall survival of 25% in phase II trials up to 2007 [1]. This changed with the introduction of anti-CTLA4 therapy in 2011 [2] and anti-PD1 therapy in 2015 [3,4]. In patients treated with anti-PD1 antibodies, real-world 1-year overall survival is now 67%, with 40% of patients achieving durable remissions of several years [5]. For patients treated with anti-PD1 plus anti-CTLA4 therapy, 5-year overall survival is reported to be as high as 52% [6].

### However, not all patients benefit from checkpoint inhibitors

At 6 months after start of anti-PD1 treatment, 43% of patients experience progression or death. Furthermore, overall survival of patients with progression at 6 months was shown to be only 16% at 30 months. This is in contrast to a 30-month overall survival of 60%, 79% and 96% for patients with stable disease, partial response and complete response at 6 months of follow-up, respectively, in real-world data [5]. Similar results were reported in patients treated with anti-PD1 plus anti-CTLA4 therapy [7].

### Accurate prediction of treatment benefit is an important topic for several reasons

First, treatment with checkpoint inhibitors is associated with severe and potentially fatal or irreversible toxicity. Severe toxicity occurs in 10-15% of patients treated with anti-PD1 monotherapy [5,8–10], and in as much as 60% of patients treated with anti-PD1 plus anti-CTLA4 combination therapy [11]. Second, checkpoint inhibition therapy is very costly. Depending on country and setting, estimates of additional costs per gained quality adjusted life year range from 25,000 to 81,000 United States Dollars [12,13]. Lastly, if patients who will not benefit are identified before start of treatment, alternative or experimental therapies can be started without delay.

### Previously identified predictors for treatment outcomes are not yet sufficient to guide clinical decisions

Known clinical predictors of poor outcome include high tumor load, presence of liver metastases and symptomatic brain metastases, increased lactate dehydrogenase (LDH) and worse Eastern Cooperative Oncology Group (ECOG) performance status [14]. In addition, other biomarkers have been explored, such as PD-L1 expression, tumor mutational burden and histopathology features. Thus far, however, these predictors are not strong enough to predict treatment outcomes with high certainty [15], or the results remain to be validated in future studies [16].

### Radiomics are by now an established modality for diagnosis, prognosis and prediction

Radiomics capture information about shape, intensity and texture of lesions in imaging and thereby form a reflection of tumor characteristics, such as necrosis or vascularization. These extracted features can subsequently be correlated to a clinical outcome [17]. This makes radiomics a cheap and non-invasive modality to, for example, discern benign from malignant lung nodules [18], estimate prognosis in non-small cell lung cancer (NSCLC) patients [19] and assess mutation status in glioblastoma [20]. Regarding prediction of checkpoint inhibitor treatment outcomes, promising findings have been published, particularly in NSCLC patients [21].

### The added value of CT radiomics for predicting durable clinical benefit to checkpoint inhibitors in melanoma remains to be determined in large multicenter studies

Three previous smaller studies have investigated radiomics for this purpose, with conflicting findings. The studies by Trebeschi et al. [22] and Peisen et al. [23] report a significant discriminative value of radiomics for treatment outcomes (AUROC=0.78 on a dataset of 80 patients, and AUROC=0.64 on a dataset of 262 patients, respectively). In contrast, Brendlin et al. [24] reported a non-discriminative performance, despite using a very similar methodology (AUROC=0.50 in 140 patients). These differences in results highlight the importance of a large dataset to determine the value of radiomics. Furthermore, only the study by Peisen et al. investigated the added value over a simpler clinical model, with varying results across different outcomes. Lastly, none of the previous studies evaluated their model on data from other centers, although variability in scanner protocol may add significant noise [25]. In this study, we aimed to address these limitations and determine the added value of radiomics for predicting checkpoint inhibitor outcomes in a multicenter study in advanced melanoma.

## Methods

### Patient selection

Eligible patients were retrospectively identified from high-quality registry data [26] from nine participating centers in The Netherlands (Amphia Ziekenhuis, Isala Zwolle, Leids Universitair Medisch Centrum, Máxima MC, Medisch Spectrum Twente, Radboudumc, UMC Utrecht, Amsterdam UMC, Zuyderland MC). Patients over the age of 18 were included if they received first-line treatment with anti-PD1 ± anti-CTLA4 checkpoint inhibition for irresectable stage IIIC or stage IV cutaneous melanoma after 01-01-2016. Exclusion criteria were (i) unavailability of baseline contrast enhanced CT imaging (CE-CT), (ii) lack of eligible target lesions, and (iii) missing follow-up data. Clinical characteristics were collected for the included patients and compared to those of the excluded patients. CT acquisition characteristics were extracted for included patients.

### Lesion selection and segmentation

For every patient, one to five lesions were selected on baseline CT imaging and segmented. We aimed to make this selection of lesions as informative and representative as possible by using the following protocol: first, the five largest lesions were selected with a maximum of two per organ. If more lesions remained after segmenting a maximum of two per organ, the largest remaining lesions were segmented up to a total of five. Lesion selections were made without knowledge of the outcome. Lesions were excluded if they were not well-demarcated, affected by imaging artifacts or if the maximum diameter was less than 5mm. Segmentations were performed in 3D Slicer [27] on the series with the lowest slice thickness by authors LSM and IAJD, under supervision of board-certified radiologists with 17 and 18 years of experience (PJ and TL, respectively).

### Feature extraction

Features were extracted from the segmented volumes using PyRadiomics [28]. For every volume, 1874 features were extracted at five different levels of detail, resulting in a total of 9370 features. An overview of the extracted features is given in the Supplementary Methods. Interobserver agreement of segmentations and features was calculated using Dice scores and intraclass correlation coefficient (ICC), respectively, based on 16 scans segmented by both observers (LSM, IAJD).

### Outcome definition

The primary outcome was durable clinical benefit, defined as a best overall response of partial or complete response, or stable disease per RECIST 1.1 [29] for a minimum of six months after start of treatment. The secondary outcome was objective response, defined as a best overall response of partial or complete response. Durable clinical benefit was used as the primary outcome, as the intended use of the model was to identify patients who would quickly progress despite treatment and therefore not derive any benefit from treatment. Individual lesion response was assessed using maximum diameter recordings at baseline and at three, six and nine months, or until treatment was changed. Given the possibility of pseudo-progression, the last available follow-up was used to determine lesion outcomes. If the maximum diameter at the last follow-up was less than 120% of the baseline diameter, the lesion was labeled as ‘does benefit’, and ‘does not benefit’ otherwise. In parallel, the lesion was labeled as ‘responsive’ if the maximum diameter was less than 70% of the baseline diameter at the last follow-up, and ‘non-responsive’ otherwise. These lesion-level cut-offs were chosen in correspondence with the patient-level cut-offs used in RECIST 1.1 [29].

### Evaluated models

Three predictive models were compared: a model based on radiomics, a model based on baseline clinical characteristics and an ensemble model that combined the predictions of these models. The radiomics model consisted of a machine learning pipeline that automatically selected optimal components and hyperparameters for feature selection, dimensionality reduction and classification (Figure 1). This pipeline was trained to predict outcomes per lesion; these outputs per lesion were then aggregated to a patient level prediction. The clinical model used the same machine learning pipeline, which was fitted on five clinical variables that were consistently shown to be predictive of checkpoint inhibitor treatment outcomes in previous literature [5,14,30,31]. These predictors were (i) ECOG performances status, (ii) LDH level, presence of (iii) brain and (iv) liver metastases, and (v) number of affected organs. All variables were one-hot encoded; missing values were encoded as a separate label. The ensemble model consisted of a logistic regression fitted on the output of the radiomics and clinical model. All three models were evaluated using a nested cross validation. The inner loop was used for optimal model selection and hyperparameter tuning; the outer loop was used to evaluate predictive performance on unseen data and was conducted in a leave-one-center-out manner. Further details are supplied in the Supplementary Methods.

**Figure 1.**
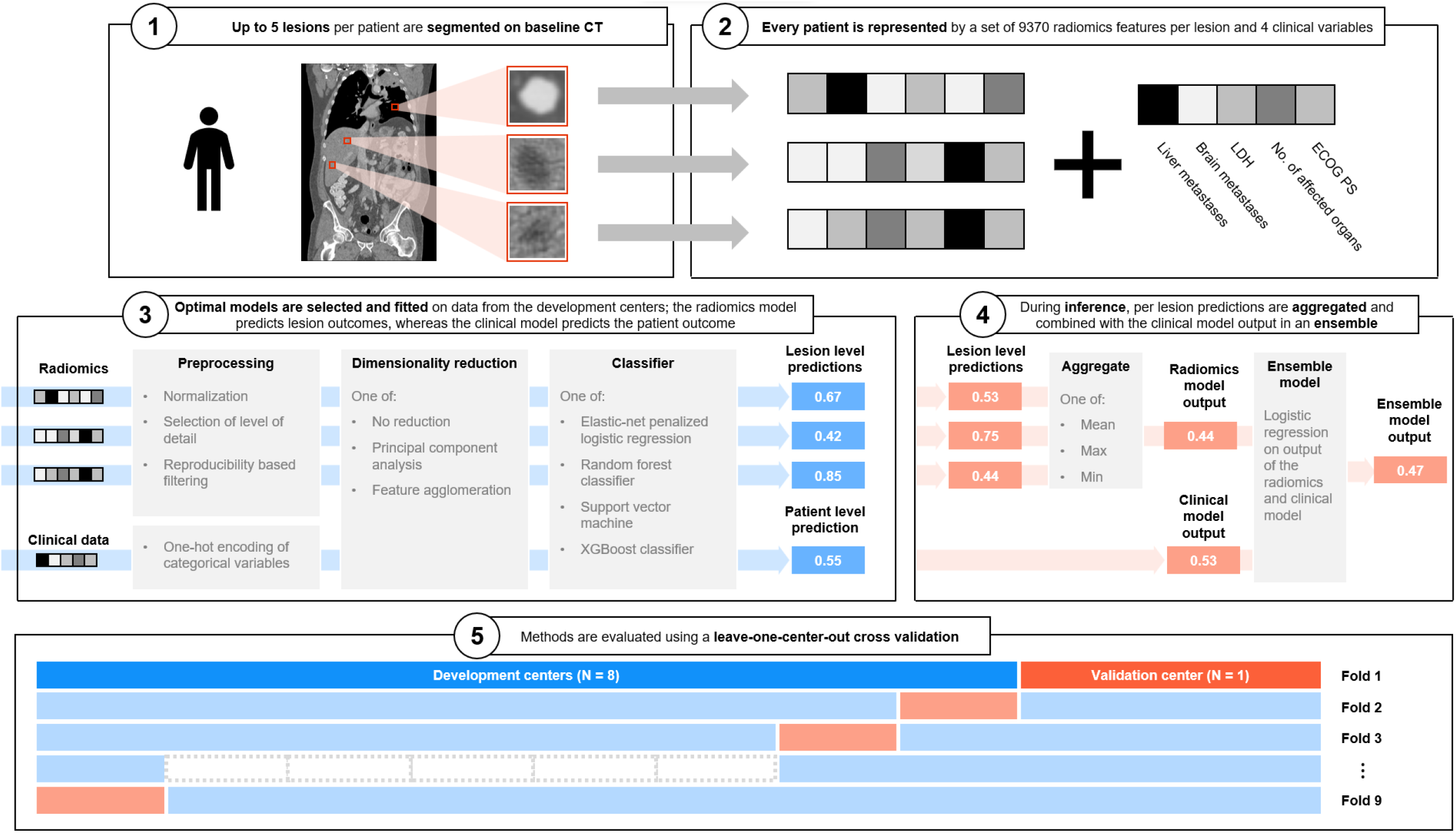
Overview of methodology.

### Statistical analysis

The discriminative performance of the models was evaluated using the area under the receiver operator characteristic curve (AUROC) and corresponding 95% confidence interval. The cross validated AUROC and confidence interval were calculated using the cvAUC R package [32]. Methods for comparing cross validated AUROCs between models are detailed in the Supplementary Methods. Subgroup analyses were conducted for patients treated with anti-PD1 therapy and anti-PD1 plus anti-CTLA4 therapy by evaluating the fitted model only on patients from the respective groups. The output of the radiomics model for predicting durable clinical benefit was correlated to the input variables of the clinical model to determine if the radiomics model learned features that were already represented in the baseline clinical model.

### Adherence to quality standards

The TRIPOD checklist [33] was completed and is available in Supplementary Table 1. The study design was reviewed by the Medical Ethics Committee and not considered subject to the Medical Research Involving Human Subjects Act in compliance with Dutch regulations; informed consent was waived.

## Results

### Patient characteristics

Out of 1191 eligible patients, 620 patients with a total of 2352 lesions were included. A flowchart of the selection process is shown in Figure 2. The rate of durable clinical benefit was 59.2% (367 patients); the objective response rate was 51.3% (318 patients); Lesion level outcomes were available for 74.4% of lesions; lesion level outcomes could not be recorded for patients from the Radboudumc (327 lesions, 13.9%). Rate of benefit was 79.4% among lesions with available labels, whereas response rate was 54.8% (Supplementary Table 5). Of all eligible patients, 490 patients were excluded because of the unavailability of contrast-enhanced pre-treatment CT. In most of these cases, an 18-fluorodeoxyglucose positron emission tomography (FDG-PET) with low-dose CT was made. Characteristics for the included patients are shown in Table 1 and compared to those of excluded patients in Supplementary Table 2. The subgroups of patient treated with anti-PD1 and combination therapy consisted of 370 and 250 patients, respectively. Supplementary Tables 3 and 4 show patient characteristics per center, and for the subgroups treated with monotherapy and combination therapy, respectively. CT acquisition characteristics per center are displayed in Supplementary Table 6.

**Figure 2.**
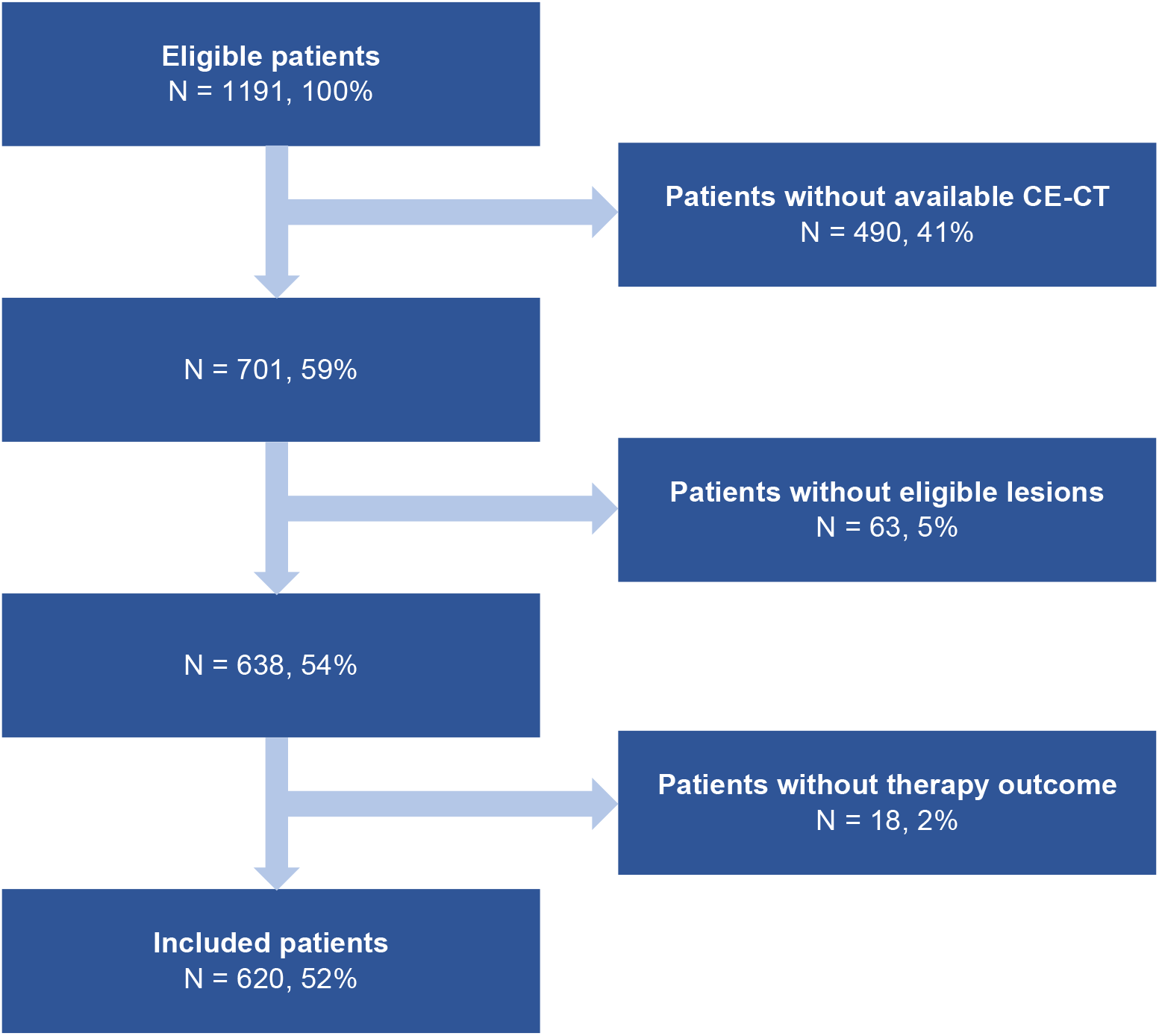
Flowchart of patient selection.

**Table 1.** Characteristics of included patients.

|  |  | **Missing** | **Overall** |
| --- | --- | --- | --- |
| **n** |  |  | 620 |
| **Age, median [Q1, Q3]** |  | 0 | 67.5 [58.0,75.0] |
| **Sex, n (%)** | Female | 0 | 239 (38.5) |
|  | Male |  | 381 (61.5) |
| **Stage, n (%)** | IIIC | 4 | 25 (4.1) |
|  | M1a |  | 49 (8.0) |
|  | M1b |  | 94 (15.3) |
|  | M1c |  | 296 (48.1) |
|  | M1d |  | 152 (24.7) |
| **ECOG performance status, n (%)** | 0 | 26 | 287 (48.3) |
|  | 1 |  | 247 (41.6) |
|  | 2-4 |  | 60 (10.1) |
| **Primary tumor location, n (%)** | Acral | 10 | 15 (2.5) |
|  | Extremity |  | 167 (27.4) |
|  | Head, neck |  | 66 (10.8) |
|  | Trunk |  | 247 (40.5) |
|  | Unknown |  | 115 (18.9) |
| **Brain metastases, n (%)** | Absent | 45 | 423 (73.6) |
|  | Asymptomatic |  | 76 (13.2) |
|  | Symptomatic |  | 76 (13.2) |
| **Liver metastases, n (%)** | Absent | 30 | 398 (67.5) |
|  | Present |  | 192 (32.5) |
| **No. of affected organs, n (%)** | <3 | 0 | 338 (54.5) |
|  | >2 |  | 282 (45.5) |
| **LDH, n (%)** | Normal | 9 | 381 (62.4) |
|  | 1-2x upper limit of normal |  | 177 (29.0) |
|  | >2x upper limit of normal |  | 53 (8.7) |
| **Durable clinical benefit, n (%)** | No benefit | 0 | 253 (40.8) |
|  | Benefit |  | 367 (59.2) |
| **Objective response, n (%)** | No response | 0 | 302 (48.7) |
|  | Response |  | 318 (51.3) |
| **Therapy, n (%)** | Anti-PD1 | 0 | 370 (59.7) |
|  | Ipilimumab & Nivolumab |  | 250 (40.3) |

### Interobserver variability

52 lesions in 16 scans were segmented by two observers. Segmentations corresponded with a median Dice score of 0.88 (IQI 0.82-0.92). For the extracted features, the median intraclass correlation coefficient was 0.97 (IQI 0.92-0.99).

### Treatment outcome prediction

For predicting durable clinical benefit, the radiomics model achieved an AUROC of 0.607 [95% CI 0.562-0.652], the clinical model an AUROC of 0.646 [95% CI 0.600-0.692], and the ensemble model an AUROC of 0.636 [95% CI 0.592-0.680]. The difference in AUROC between the ensemble and clinical model was not statistically significant (Supplementary Figure 1). Calibration curves showed adequate calibration of the three models with no evidence of poor fit (Hosmer-Lemeshow p > 0.07). The range of predicted probabilities was comparable between models (IQI 0.56-0.65, 0.53-0.67 and 0.52-0.69 for the radiomics, clinical and ensemble model, respectively). Results were similar for predicting objective response (Supplementary Figure 2-3). Predictive performance for both outcomes was comparable in subgroups of patients treated with monotherapy and combination therapy, with a trend of better discrimination in the subgroup of patients treated with combination therapy (Supplementary Figures 4-7). Details of the selected models and hyperparameters per fold are shown in Supplementary Table 7.

### Comparison of radiomics and clinical model

The predicted probability of durable clinical benefit by the radiomics model was significantly lower in patients in whom liver metastases were absent (Mann-Whitney U, p < 0.001, Figure 4A), in patients with higher LDH (Kruskal-Wallis p < 0.001, Figure 4D) and who had more affected organs (Mann-Whitney U p < 0.001, Figure 4E). The output of radiomics model was not significantly different in patients with and without brain metastases), and for different categories of ECOG performance status (Figure 4B-C). The output of radiomics and clinical models were significantly and positively correlated (Spearman’ s correlation coefficient = 0.369, p < 0.001, Figure 4F).

**Figure 3.**
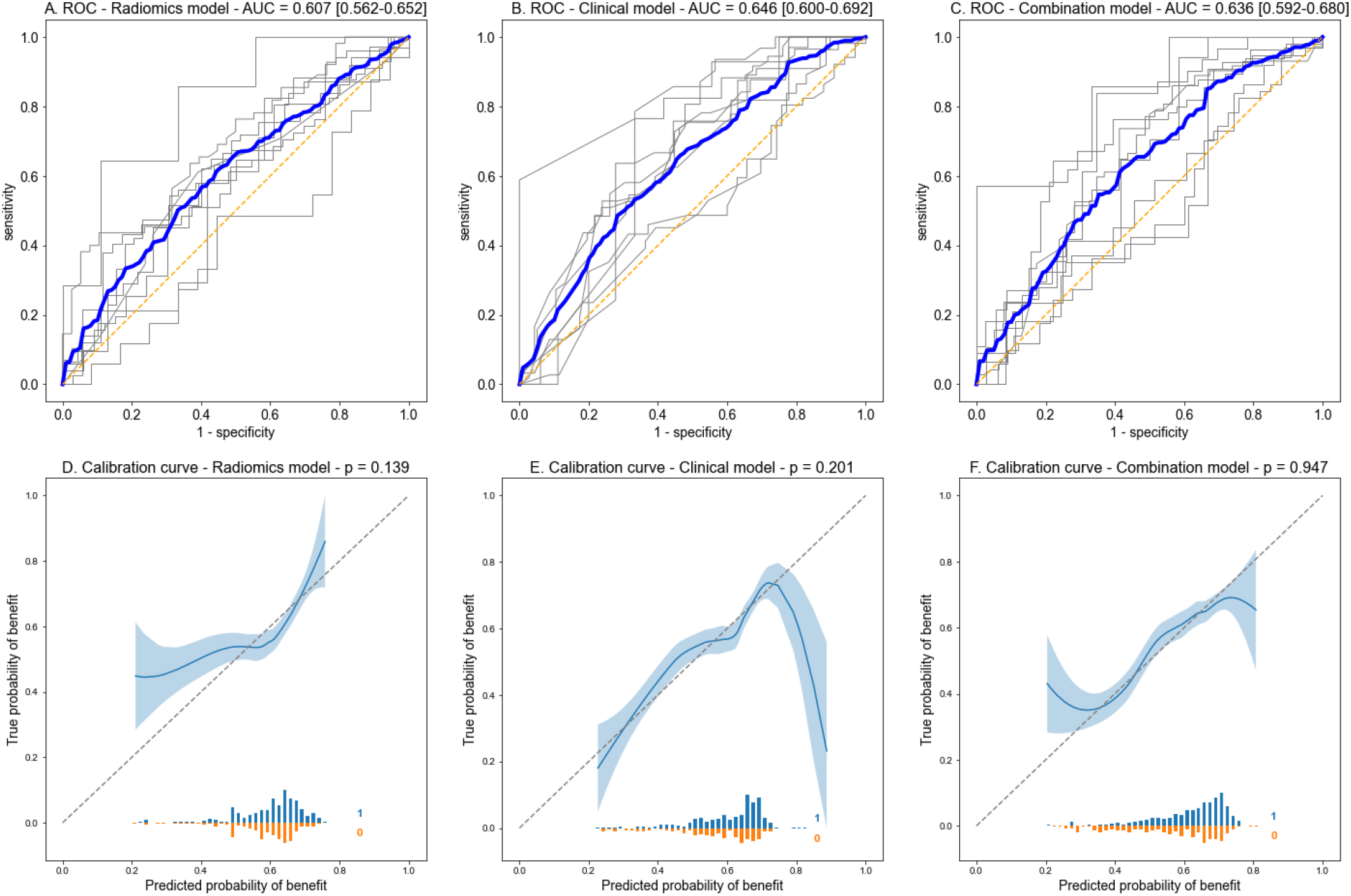
Receiver operator characteristic curves and calibration curves for predicting durable clinical benefit. (A-C) Receiver operator characteristic (ROC) curves for predicting durable clinical benefit in patients with melanoma treated with anti-PD1 ± anti-CTLA4 checkpoint inhibition for the radiomics model (A), clinical model (B) and combination model (C). Gray curves correspond to results per fold; blue curves are the weighted average of the results per fold. The area under the curve (AUC) with corresponding 95% confidence intervals are displayed. (D-F) LOESS fitted calibration curves for predicting durable clinical benefit in the radiomics model (D), clinical model (E) and combination model (F); the shaded area corresponds to ±1 standard deviation. Histograms of the predictions for positive (blue) and negative (orange) samples are provided below the curves, the x-axis displays the predicted values for these histograms. P-values of the Hosmer-Lemeshow goodness-of-fit test are shown in the plot titles.

**Figure 4.**
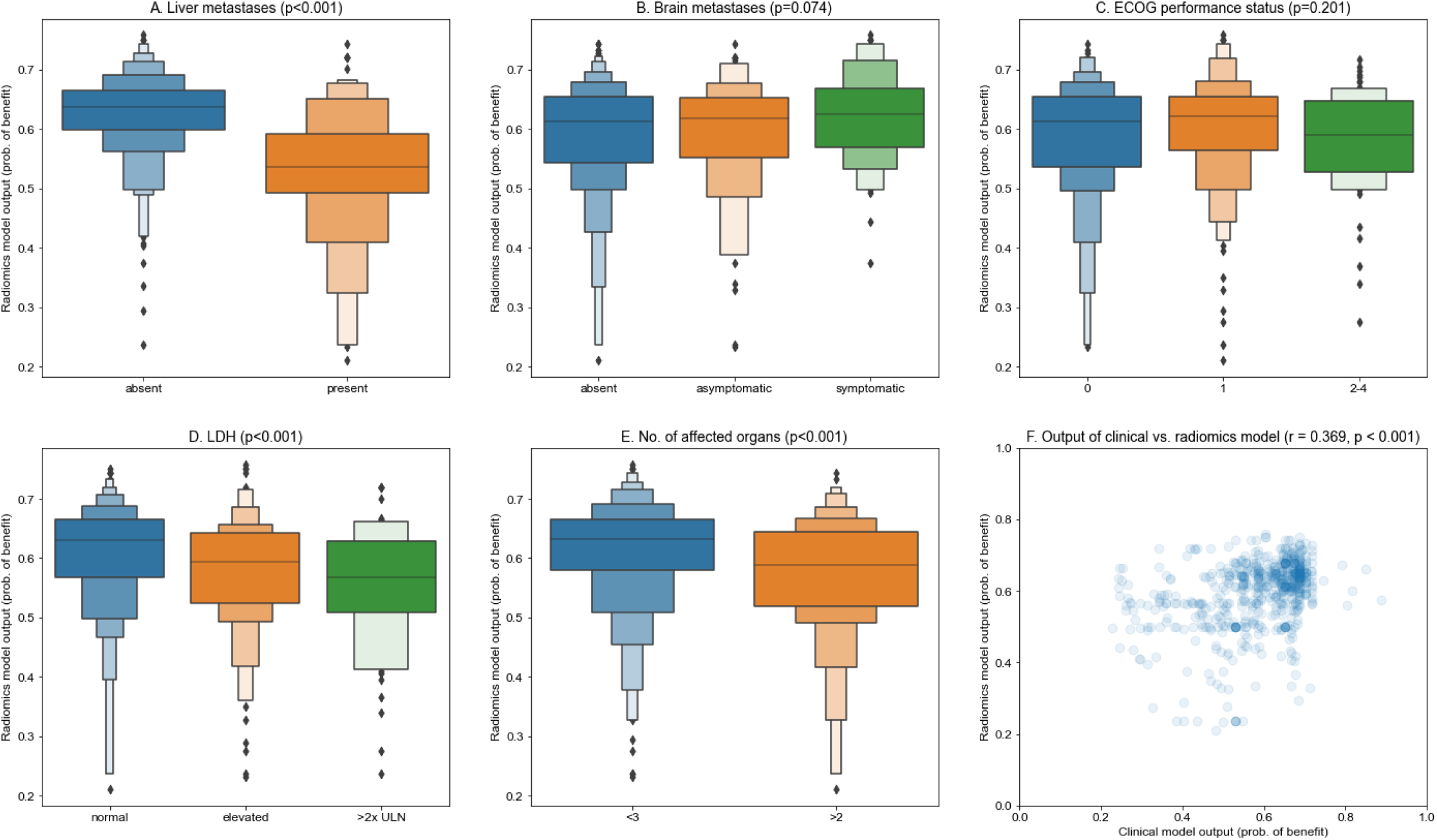
Correspondence between the output of the clinical and radiomics models for predicting durable clinical benefit. Graphical overview of correspondence between the output of the radiomics and clinical models. (A-E) Boxenplots of the output of the radiomics model, compared across different values for clinical predictors. (A) The output of the radiomics model is significantly lower in patients with liver metastases than in patients without (Mann-Whitney U p < 0.001). (B) No statistical difference was found in the output of the radiomics model between patients without or with asymptomatic or symptomatic brain metastases (Kruskal-Wallis p = 0.074) and ECOG performance status (Kruskal-Wallis p = 0.201). D) The output of the radiomics model is significantly lower in patients with higher levels of LDH (Kruskal-Wallis p < 0.001) and with more affected organs (Mann-Whitney U p < 0.001). (F) The outputs of the clinical and radiomics models (predicted probability of response) are positively correlated (Spearman’ s rank correlation coefficient = 0.369, p < 0.001).

## Discussion

### Overview

#### The present work shows that radiomics are moderately predictive of checkpoint inhibitor treatment outcomes in patients with advanced melanoma

The results were consistent for both durable clinical benefit and objective response rate, and are most in line with the findings of the earlier study by Peisen et al. A recent work by Dercle et al. allows for comparison to a model that also incorporates radiomics from on-treatment CT scans [34]. This model reached an AUROC of 0.92 for predicting overall survival at six months, indicating that on-treatment radiomics are strongly predictive. However, most toxicity occurs in the first three months and long-term outcomes can already be accurately predicted using on-treatment information without the use of radiomics [6,35]. Predicting response using the 3-month on-treatment scan therefore appears to be of limited clinical relevance.

#### Addition of radiomics to known clinical predictors, however, did not yield improvement in predictive value

The combination model was not superior to the clinical model in either discrimination or calibration. This lack of improvement can be explained due to an overlap in the information learned by the radiomics model, and the information that is already represented in clinical variables. As demonstrated, the radiomics model indirectly learns to detect the presence of liver metastases and the amount of tumor burden, as reflected in LDH and number of affected organs. The fact that this information is indeed learned as expected is a strong argument for the validity of the present work. Furthermore, this indicates that such overlap is likely to be present in any radiomics model which is investigated for clinical purposes.

### Future research

#### Studies on radiomics should assess the added value over simpler predictors

Many smaller exploratory studies have been conducted into the predictive value of radiomics for checkpoint inhibitor outcomes across different malignancies [21]. Their findings are almost exclusively positive, but the added value over clinical predictors was seldomly assessed. The present work demonstrates that clinical predictors can be captured by radiomics, and that the added value of radiomics should therefore always be investigated, even in exploratory studies.

#### Future works should aim to improve on radiomics through deep learning or spectral CT derived radiomics

Deep learning has a significant advantage over handcrafted radiomics, as this method is not limited by predefined features in what information can be captured. Instead, a deep learning approach is given the raw data as input and learns informative features on the fly [36]. Furthermore, spectral CT derived radiomics were shown to be superior over single energy radiomics for predicting response to checkpoint inhibition in patients with melanoma by Brendlin et al. [24]. As spectral CT scanners become increasingly available, this approach may be tested more thoroughly in future research.

#### Lastly, the multimodal approach should be extended with other data sources

Accurately predicting checkpoint inhibitor treatment outcomes in melanoma remains challenging. It is possible that individual biomarkers are insufficient to guide clinical decisions. An approach that combines different data sources may therefore prove to be superior. A possible modalities that may be explored for this purpose is histopathology imaging [16], which will be investigated in this cohort in a future work.

### Strengths and limitations

#### The strengths of this work are the large sample size, the multicenter design and extensive hyperparameter optimization

This is the largest work published on radiomics for prediction of checkpoint inhibitor treatment outcomes in any malignancy [21]. This large size adds to the weight of the presented conclusion. Furthermore, the dataset in this work includes patients from nine different centers. As stability of radiomics features across scanner types and protocols is far from certain, external validation is essential for determining the practical value of a radiomics approach. Lastly, the proposed pipeline systematically explores design choices, from the extraction of radiomics to the final prediction. This approach should maximize potential performance by avoiding arbitrary and therefore possibly suboptimal design choices.

#### A potential limitation is the exclusion of a large fraction of patients due to unavailability of CE-CT imaging

Comparison of patient characteristics between the included and excluded groups showed minor differences overall, with a trend towards more progressed disease in the included patients. Our hypothesis for this is that patients with more progressed disease are more likely to directly present to medical oncology, instead of being referred after having undergone imaging by a different specialty where FDG-PET is the preferred modality. Although this selection may theoretically have influenced the presented results, this risk is arguably limited as the characteristics of in- and excluded patients are overall very comparable.

## Conclusion

### In conclusion, radiomics are predictive of checkpoint inhibition treatment outcomes in patients with advanced melanoma, but offer limited added value over a simpler clinical model

A radiomics model can predict both durable clinical benefit and response from checkpoint inhibitor therapy with moderate discriminative performance. However, the predictive value of this radiomics model overlaps with that of a clinical model, which is evident from the lack of improvement of a combined model. The added value of a radiomics approach therefore appears to be limited. Future research should focus on related techniques, such as deep learning or radiomics on dual energy CT images. In addition, an approach that combines radiomics and clinical data with other modalities may provide a next step towards accurate prediction of checkpoint inhibitor treatment outcomes in melanoma.

## Data Availability

N/A

## Conflict of interest statement

AvdE has advisory relationships with Amgen, Bristol Myers Squibb, Roche, Novartis, MSD, Pierre Fabre, Sanofi, Pfizer, Ipsen, Merck and has received research study grants not related to this paper from Sanofi, Roche, Bristol Myers Squibb, Idera and TEVA and has received travel expenses from MSD Oncology, Roche, Pfizer and Sanofi and has received speaker honoraria from BMS and Novartis.

JdG has consultancy/advisory relationships with Bristol Myers Squibb, Pierre Fabre, Servier, MSD, Novartis.

PJ has a research collaboration with Philips Healthcare and Vifor Pharma.

MBS has consultancy/advisory relationships with Pierre Fabre, MSD and Novartis.

EK has consultancy/advisory relationships with Bristol Myers Squibb, Novartis, Merck, Pierre Fabre, Lilly, Bayer, EISAI and Ipsen, and received research grants not related to this paper from Bristol Myers Squibb and Pierre Fabre.

PD has consultancy/advisory relationships with Paige, Pantarei and Samantree paid to the institution and research grants from Pfizer, none related to current work and paid to institute.

KS has advisory relationships with Bristol Myers Squibb, Novartis, MSD, Pierre Fabre, AbbVie and received honoraria from Novartis, MSD and Roche and research funding from Bristol Myers Squibb, TigaTx and Philips.

TL has received research funding from Philips.

All remaining authors have declared no conflicts of interest.

