## Supplementary files for "CT radiomics to predict checkpoint inhibitors treatment outcomes in patients with advanced cutaneous melanoma"

### Supplementary Methods

#### Radiomics extraction

The version used for PyRadiomics was v3.0.1. For every lesion, the radiomics feature extraction was conducted at five levels of detail (from fine to coarse):

1. Bin width = 1 Hounsfield Unit (HU), isometric resampling at 1x1x1 $mm^3$
2. Bin width = 3 HU, isometric resampling at 2x2x2 $mm^3$
3. Bin width = 5 HU, isometric resampling at 3x3x3 $mm^3$
4. Bin width = 15 HU, isometric resampling at 4x4x4 $mm^3$
5. Bin width = 25 HU, isometric resampling at 5x5x5 $mm^3$

For some lesions, features could not be extracted at the coarser levels of detail. In these lesions, the isometric resampling at larger voxel sizes resulted in only a single remaining voxel, from which features are not defined. For lesions where this was the case, the missing levels of detail were imputed with the coarsest level of detail that was available.

All features were used, computed on the original images as well as on derived images, obtained by applying all processing and filtering options. For the Laplacian of Gaussian filter, the sigma parameter was set to 1, 2.5, 5, 7.5 and 10 mm.

#### Clinical variables

The clinical model consisted of five categorical variables. The following categories were used:

- Liver metastases: absent vs. present vs. missing
- Brain metastases: absent vs. asymptomatic vs. symptomatic vs. missing
- ECOG performance status: 0 vs. 1 vs. ≥2 vs. missing
- LDH: normal vs. elevated vs. above twice the upper limit of normal vs. missing
- Number of affected organs: <3 or >2

Continuous values were not available for LDH; categorization was therefore necessary for this variable.

#### Radiomics model

All models were implemented using sci-kit learn v0.23.2 and XGBoost for Python v1.6.2. The radiomics model consisted of the following components:

1. **Feature normalization** to zero mean and unit variance.
2. **Correlation threshold**: the interobserver correlation is precomputed for every feature. In this step, all features below a certain threshold of correlation are dropped.
3. **Level of detail selection**: only features of a single level of detail are selected; all other features are dropped.
4. **Dimensionality reduction** through one of the following:
    a. No reduction (identity function)
    b. Principal component analysis
    c. Hierarchical feature agglomeration
5. **Classification** through one of the following:
    a. Elastic-net regularized logistic regression
    b. Random forest classifier
    c. Support vector machine
    d. XGBoost classifier
6. **Aggregation** of lesion level prediction to a patient level prediction through taking the mean, maximum or minimum of all lesion level predictions.

In steps 1 through 4, the radiomics features were preprocessed. In step 5, the classifier was fit to predict lesion level outcomes based on the preprocessed radiomics features. In step 6, the output per lesion was aggregated to arrive at the patient level prediction.

This six-step pipeline was optimized and tested using nested cross validation. A nested cross validation consists of an inner and outer loop. The purpose of the inner loop is to select the optimal combination of components and hyperparameters. This is done by trying many different combinations of components and hyperparameters and evaluating how well each configuration performs using the inner cross validation. The best performing model is then selected, refitted on all data used in the inner loop and then evaluated on the test set of the outer loop.

As lesion level labels are required to fit the classifier in step 5, the radiomics model only made use of lesions for which labels were available for fitting. These lesions could be used, however, during model evaluation in both the inner and outer loop, provided that the patient level outcome was available. This was the case for all patients included from the Radboud center: as lesion level labels were not available from this center, these patients were never used for model fitting. They were, however, used for model selection and evaluated once when the Radboud center was used as the test set in the outer loop of the nested cross validation.

The three dimensionality reduction methods and four classifiers yielded a total of 12 possible configurations; method for aggregation was optimized as a hyperparameter. For each of these 12 configurations, hyperparameters were explored using Bayesian optimization. Bayesian optimization is an algorithm to efficiently find the maximum of an objective function, given a set of hyperparameters [1]. In this case, the objective function was the performance of a configuration given the hyperparameters. Specifically, the performance of the model was quantified as the mean AUROC of a 5-fold inner cross validation for predicting the patient level outcome (objective response or durable clinical benefit); lesions from a single patient were grouped during the cross validation to prevent data leaking.

The Optuna v3.0.2 package was used to implement the Bayesian optimization; the Tree-structured Parzen Estimator sampler was used to sample the space of hyperparameters. Per configuration, 50 iterations were performed. The hyperparameters per component, explored ranges and scale are specified in the table below:

| **Component** | **Hyperparameter** | **Range** | **Scale** |
|---|---|---|---|
| Correlation threshold | Threshold | 0 – 0.99 | Float, uniform |
| Level of detail selector | Level of detail | [1,2,3,4,5] | Categorical |
| Principal component analysis | Number of components | 1 – 20 | Integer, uniform |
| Hierarchical feature agglomeration | Number of clusters | 1 – 20 | Integer, uniform |
| Elastic-net regularized logistic regression | Alpha | $10^{-10}$ – 1 | Float, uniform |
| | L1-ratio | 0 – 1 | Float, uniform |
| Random forest classifier | Max features | 1 – 20 | Integer, uniform |
| | Max depth | 1 – 20 | Integer, uniform |
| | Number of estimators | 10 – 1000 | Integer, uniform |
| Supper vector machine | C | $10^{-3}$ – $10^{2}$ | Float, logarithmic |
| | Gamma | $10^{-6}$ – 10 | Float, logarithmic |
| XGB Classifier | Learning rate | 0 – 1 | Float, uniform |
| | Gamma | 0 – 5 | Float, uniform |
| | Number of estimators | 1 – 100 | Integer, uniform |
| | Max depth | 1 – 20 | Integer, uniform |
| | Alpha | 0 – 5 | Float, uniform |
| | Lambda | 0 – 5 | Float uniform |
| Aggregation | Method | [Mean, Max, Min] | Categorical |

After hyperparameter optimization was complete for all 12 configurations, the configuration with the highest AUROC was selected. This configuration was then retrained on all training data in the current fold.

Lastly, the radiomics model is recalibrated to account for any miscalibration that may arise from the method of aggregating lesion level predictions to a patient level. For example, when the 'minimum method' is used for aggregation, the radiomics model may be expected to output a prediction which is too low on average. To counter this potential miscalibration, the following steps are taken:

1. The optimal radiomics model is used to make a patient level prediction for every patient in the current inner loop. To make these predictions independent, these predictions are made in a 5-fold cross validation; lesions of a single patient were grouped to prevent data leaking.
2. An unpenalized logistic regression was fitted using these independent predictions as input and the patient level labels as output. This second-stage classifier effectively recalibrates the model.

##### Clinical model

The clinical model consisted of the following steps:

1. **One-hot encoding**: categorical variables were split into multiple binary variables, resulting in a total of 17 variables; missing values were separately encoded.
2. **Dimensionality reduction:** same as in radiomics model.
3. **Classifier:** same as in radiomics model.

For the clinical model, the same hyperparameter tuning and model selection method was used as for the radiomics model. The main difference is that the clinical model is fitted directly to predict the patient level outcome, as the input variables were also at a patient level. An aggregation component was therefore not used in the clinical model.

##### Ensemble model and radiomics model recalibration

After selection of the optimal radiomics and clinical model on the data of the current training folds, the ensemble model is constructed

This is done per the following steps:

1. In the same fashion as for recalibration of the radiomics model, the optimal radiomics model is used to make a patient level prediction for every patient in the current inner loop. To make these predictions independent, these predictions are made in a 5-fold cross validation; lesions of a single patient were grouped to prevent data leaking.
2. Using the same process, independent predictions were obtained for the clinical model.
3. An unpenalized logistic regression was then fitted using the output of the radiomics and clinical models as input, and the patient level outcome as a target.

##### Inference

After optimizing and fitting the radiomics, clinical and ensemble model in the inner loop, the three models are evaluated in the outer loop. This is performed as follows:

1. The fitted radiomics model predicts lesion level outcomes based on the radiomics of every lesion in step 1 through 5; these lesion level predictions are then combined using the aggregation component in step 6.
2. The fitted clinical model directly predicts patient level outcomes based on the clinical characteristics.
3. The ensemble model combines the output of the radiomics and clinical model using the weights fitted on the data in the training folds.

##### Calculation of cross-validated AUROC

The cvAUC R package is used to calculate the cross validated AUROC and corresponding 95% confidence interval. This method uses influence curves to estimate variance of the point estimate, which is calculated as the unweighted average of the AUROC across the folds. No consensus exists, however, as to if and how AUROCs across folds should be weighted [2]. Alternatives include weighing according to center size, number of events or inverse variance. The influence of this choice on the eventual results is investigated and shown in Supplementary Table 8.

##### Comparison of AUROCs between clinical and combination model

The following procedure was used to determine statistical significance of a difference in AUROC between the investigated combination model and baseline clinical model. In every validation fold (consisting of patient from a single center), 1000 bootstrapped samples were drawn. In every bootstrapped sample, the difference in AUROC between the combination and clinical model was calculated. Per center, the mean and standard deviation of differences in AUROCs across all bootstrapped samples was recorded. Next, a random effects model was used to pool the results. Statistical significance of a difference in AUROC was defined as exclusion of zero from the 95% confidence interval of the aggregated difference in AUROC across all centers.

This method was used in absence of an alternative to the DeLong's test [3] for comparing two AUROCs in a cross validation setup. It must be noted, however, that this method does not account for the dependence of samples between folds. This has been shown to lead to estimates of variance which are too small on average [4], which implies that the resulting confidence interval of this analysis are, on average, too small.

#### Supplementary Table 1 - TRIPOD checklist

| Section/Topic | | | Checklist Item | Page |
|---|---|---|---|---|
| **Title and abstract** | | | | |
| Title | | | Identify the study as developing and/or validating a multivariable prediction model, the target population, and the outcome to be predicted. | 1 |
| Abstract | | | Provide a summary of objectives, study design, setting, participants, sample size, predictors, outcome, statistical analysis, results, and conclusions. | 1 |
| **Introduction** | | | | |
| Background and objectives | | | Explain the medical context (including whether diagnostic or prognostic) and rationale for developing or validating the multivariable prediction model, including references to existing models. | 2 |
| | | | Specify the objectives, including whether the study describes the development or validation of the model or both. | 2 |
| **Methods** | | | | |
| Source of data | | | Describe the study design or source of data (e.g., randomized trial, cohort, or registry data), separately for the development and validation data sets, if applicable. | 3 |
| | | | Specify the key study dates, including start of accrual; end of accrual; and, if applicable, end of follow-up. | 3 |
| Participants | | | Specify key elements of the study setting (e.g., primary care, secondary care, general population) including number and location of centres. | 3 |
| | | | Describe eligibility criteria for participants. | 3 |
| | | | Give details of treatments received, if relevant. | 3 |
| Outcome | | | Clearly define the outcome that is predicted by the prediction model, including how and when assessed. | 3 |
| | | | Report any actions to blind assessment of the outcome to be predicted. | N/A |
| Predictors | | | Clearly define all predictors used in developing or validating the multivariable prediction model, including how and when they were measured. | 3, supplements |
| | | | Report any actions to blind assessment of predictors for the outcome and other predictors. | 4 |
| Sample size | | | Explain how the study size was arrived at. | N/A |
| Missing data | | | Describe how missing data were handled (e.g., complete-case analysis, single imputation, multiple imputation) with details of any imputation method. | Supplements |
| Statistical analysis methods | | | Describe how predictors were handled in the analyses. | 3-4, supplements |
| | | | Specify type of model, all model-building procedures (including any predictor selection), and method for internal validation. | 3-4, supplements |
| | | | For validation, describe how the predictions were calculated. | 3-4, supplements |
| | | | Specify all measures used to assess model performance and, if relevant, to compare multiple models. | 4 |
| | | | Describe any model updating (e.g., recalibration) arising from the validation, if done. | N/A |
| Risk groups | | | Provide details on how risk groups were created, if done. | N/A |
| Development vs. validation | | | For validation, identify any differences from the development data in setting, eligibility criteria, outcome, and predictors. | 3 |
| **Results** | | | | |
| Participants | | | Describe the flow of participants through the study, including the number of participants with and without the outcome and, if applicable, a summary of the follow-up time. A diagram may be helpful. | 4, figure 2 |
| | | | Describe the characteristics of the participants (basic demographics, clinical features, available predictors), including the number of participants with missing data for predictors and outcome. | Table 1, figure 2 |
| | | | For validation, show a comparison with the development data of the distribution of important variables (demographics, predictors and outcome). | Supplements |
| Model development | | | Specify the number of participants and outcome events in each analysis. | Table 1, figure 2 |
| | | | If done, report the unadjusted association between each candidate predictor and outcome. | N/A |
| Model specification | | | Present the full prediction model to allow predictions for individuals (i.e., all regression coefficients, and model intercept or baseline survival at a given time point). | N/A |
| | | | Explain how to the use the prediction model. | N/A |
| Model performance | | | Report performance measures (with CIs) for the prediction model. | 4, figure 3 |
| Model-updating | | | If done, report the results from any model updating (i.e., model specification, model performance). | N/A |
| **Discussion** | | | | |
| Limitations | | | Discuss any limitations of the study (such as nonrepresentative sample, few events per predictor, missing data). | 5-6 |
| Interpretation | | | For validation, discuss the results with reference to performance in the development data, and any other validation data. | N/A |
| | | | Give an overall interpretation of the results, considering objectives, limitations, results from similar studies, and other relevant evidence. | 5 |
| Implications | | | Discuss the potential clinical use of the model and implications for future research. | 5-6 |
| **Other information** | | | | |
| Supplementary information | | | Provide information about the availability of supplementary resources, such as study protocol, Web calculator, and data sets. | N/A |
| Funding | | | Give the source of funding and the role of the funders for the present study. | 7 |

#### Supplementary Table 2 - Characteristics of included and excluded patients

| | | Missing | Overall | Excluded | Included |
|---|---|---|---|---|---|
| **n** | | | 1191 | 571 | 620 |
| **Age, median [Q1, Q3]** | | 0 | 67.0 [57.0,75.0] | 67.0 [56.0,75.5] | 67.5 [58.0,75.0] |
| **Sex, n (%)** | Female | 0 | 445 (37.4) | 206 (36.1) | 239 (38.5) |
| | Male | | 746 (62.6) | 365 (63.9) | 381 (61.5) |
| **Stage, n (%)** | IIIC | 9 | 93 (7.9) | 68 (12.0) | 25 (4.1) |
| | M1a | | 100 (8.5) | 51 (9.0) | 49 (8.0) |
| | M1b | | 163 (13.8) | 69 (12.2) | 94 (15.3) |
| | M1c | | 534 (45.2) | 238 (42.0) | 296 (48.1) |
| | M1d | | 292 (24.7) | 140 (24.7) | 152 (24.7) |
| **ECOG performance status, n (%)** | 0 | 53 | 536 (47.1) | 249 (45.8) | 287 (48.3) |
| | 1 | | 505 (44.4) | 258 (47.4) | 247 (41.6) |
| | 2-4 | | 97 (8.5) | 37 (6.8) | 60 (10.1) |
| **Primary tumor location, n (%)** | Acral | 17 | 25 (2.1) | 10 (1.8) | 15 (2.5) |
| | Extremity | | 362 (30.8) | 195 (34.6) | 167 (27.4) |
| | Head, neck | | 152 (12.9) | 86 (15.2) | 66 (10.8) |
| | Trunk | | 443 (37.7) | 196 (34.8) | 247 (40.5) |
| | Unknown | | 192 (16.4) | 77 (13.7) | 115 (18.9) |
| **Brain metastases, n (%)** | Absent | 137 | 762 (72.3) | 339 (70.8) | 423 (73.6) |
| | Asymptomatic | | 162 (15.4) | 86 (18.0) | 76 (13.2) |
| | Symptomatic | | 130 (12.3) | 54 (11.3) | 76 (13.2) |
| **Liver metastases, n (%)** | Absent | 108 | 774 (71.5) | 376 (76.3) | 398 (67.5) |
| | Present | | 309 (28.5) | 117 (23.7) | 192 (32.5) |
| **No. of affected organs, n (%)** | <3 | 0 | 682 (57.3) | 344 (60.2) | 338 (54.5) |
| | >2 | | 509 (42.7) | 227 (39.8) | 282 (45.5) |
| **LDH, n (%)** | Normal | 17 | 767 (65.3) | 386 (68.6) | 381 (62.4) |
| | 1-2x upper limit of normal | | 318 (27.1) | 141 (25.0) | 177 (29.0) |
| | >2x upper limit of normal | | 89 (7.6) | 36 (6.4) | 53 (8.7) |
| **Durable clinical benefit, n (%)** | No benefit | 62 | 445 (39.4) | 192 (37.7) | 253 (40.8) |
| | Benefit | | 684 (60.6) | 317 (62.3) | 367 (59.2) |
| **Response, n (%)** | No response | 62 | 531 (47.0) | 229 (45.0) | 302 (48.7) |
| | Response | | 598 (53.0) | 280 (55.0) | 318 (51.3) |
| **Therapy, n (%)** | Anti-PD1 | 0 | 750 (63.0) | 380 (66.5) | 370 (59.7) |
| | Ipilimumab & Nivolumab | | 441 (37.0) | 191 (33.5) | 250 (40.3) |

Abbreviations – ECOG: Eastern Cooperative Oncology Group; LDH: lactate dehydrogenase.

#### Supplementary Table 3 – Patient characteristics per center

| | | Missing | Overall | Amphia | Isala | LUMC | Maxima | MST | Radboud | UMCU | AUMC | Zuyderland |
|---|---|---|---|---|---|---|---|---|---|---|---|---|
| **n** | | | 620 | 51 | 96 | 72 | 54 | 23 | 84 | 93 | 118 | 29 |
| **Age, median [Q1,Q3]** | | 0 | 67.5 [58.0,75.0] | 65.0 [54.0,71.0] | 69.0 [59.0,76.0] | 71.0 [62.0,77.2] | 66.0 [58.5,75.0] | 63.0 [53.5,69.5] | 63.0 [54.0,71.0] | 69.0 [58.0,74.0] | 70.0 [59.0,76.0] | 66.0 [55.0,71.0] |
| **Sex, n (%)** | Female | 0 | 239 (38.5) | 26 (51.0) | 34 (35.4) | 23 (31.9) | 19 (35.2) | 6 (26.1) | 33 (39.3) | 33 (35.5) | 53 (44.9) | 12 (41.4) |
| | Male | | 381 (61.5) | 25 (49.0) | 62 (64.6) | 49 (68.1) | 35 (64.8) | 17 (73.9) | 51 (60.7) | 60 (64.5) | 65 (55.1) | 17 (58.6) |
| **Stage, n (%)** | IIIC | 4 | 25 (4.1) | 1 (2.0) | 5 (5.2) | 1 (1.4) | 2 (3.8) | 2 (8.7) | 1 (1.2) | 6 (6.5) | 7 (6.0) | |
| | M1a | | 49 (8.0) | 7 (13.7) | 10 (10.4) | 5 (7.0) | 6 (11.3) | 2 (8.7) | 7 (8.4) | 2 (2.2) | 5 (4.3) | 5 (17.2) |
| | M1b | | 94 (15.3) | 7 (13.7) | 18 (18.8) | 9 (12.7) | 7 (13.2) | 4 (17.4) | 16 (19.3) | 15 (16.1) | 14 (12.0) | 4 (13.8) |
| | M1c | | 296 (48.1) | 24 (47.1) | 41 (42.7) | 34 (47.9) | 23 (43.4) | 9 (39.1) | 41 (49.4) | 38 (40.9) | 70 (59.8) | 16 (55.2) |
| | M1d | | 152 (24.7) | 12 (23.5) | 22 (22.9) | 22 (31.0) | 15 (28.3) | 6 (26.1) | 18 (21.7) | 32 (34.4) | 21 (17.9) | 4 (13.8) |
| **ECOG performance status, n (%)** | 0 | 26 | 287 (48.3) | 27 (56.2) | 68 (72.3) | 27 (41.5) | 44 (84.6) | 5 (23.8) | 22 (26.5) | 21 (23.9) | 61 (52.1) | 12 (46.2) |
| | 1 | | 247 (41.6) | 20 (41.7) | 15 (16.0) | 35 (53.8) | 4 (7.7) | 12 (57.1) | 55 (66.3) | 45 (51.1) | 47 (40.2) | 14 (53.8) |
| | 2-4 | | 60 (10.1) | 1 (2.1) | 11 (11.7) | 3 (4.6) | 4 (7.7) | 4 (19.0) | 6 (7.2) | 22 (25.0) | 9 (7.7) | |
| **Primary tumor location, n (%)** | Acral | 10 | 15 (2.5) | 3 (5.9) | 4 (4.2) | | 3 (5.6) | | 1 (1.2) | 2 (2.2) | 2 (1.8) | |
| | Extremity | | 167 (27.4) | 10 (19.6) | 23 (24.2) | 20 (28.6) | 12 (22.2) | 4 (17.4) | 25 (29.8) | 23 (25.6) | 41 (36.0) | 9 (31.0) |
| | Head, neck | | 66 (10.8) | 7 (13.7) | 6 (6.3) | 10 (14.3) | 7 (13.0) | 2 (8.7) | 13 (15.5) | 6 (6.7) | 12 (10.5) | 3 (10.3) |
| | Trunk | | 247 (40.5) | 22 (43.1) | 37 (38.9) | 24 (34.3) | 22 (40.7) | 12 (52.2) | 29 (34.5) | 42 (46.7) | 45 (39.5) | 14 (48.3) |
| | Unknown | | 115 (18.9) | 9 (17.6) | 25 (26.3) | 16 (22.9) | 10 (18.5) | 5 (21.7) | 16 (19.0) | 17 (18.9) | 14 (12.3) | 3 (10.3) |
| **Brain metastases, n (%)** | Absent | 45 | 423 (73.6) | 37 (75.5) | 67 (75.3) | 46 (67.6) | 28 (65.1) | 15 (71.4) | 63 (77.8) | 55 (63.2) | 87 (80.6) | 25 (86.2) |
| | Asymptomatic | | 76 (13.2) | 9 (18.4) | 11 (12.4) | 10 (14.7) | 4 (9.3) | 4 (19.0) | 10 (12.3) | 14 (16.1) | 12 (11.1) | 2 (6.9) |
| | Symptomatic | | 76 (13.2) | 3 (6.1) | 11 (12.4) | 12 (17.6) | 11 (25.6) | 2 (9.5) | 8 (9.9) | 18 (20.7) | 9 (8.3) | 2 (6.9) |
| **Liver metastases, n (%)** | Absent | 30 | 398 (67.5) | 39 (78.0) | 63 (69.2) | 51 (73.9) | 34 (65.4) | 14 (70.0) | 54 (65.1) | 64 (73.6) | 59 (54.1) | 20 (69.0) |
| | Present | | 192 (32.5) | 11 (22.0) | 28 (30.8) | 18 (26.1) | 18 (34.6) | 6 (30.0) | 29 (34.9) | 23 (26.4) | 50 (45.9) | 9 (31.0) |
| **No. of affected organs, n (%)** | <3 | 0 | 338 (54.5) | 30 (58.8) | 60 (62.5) | 36 (50.0) | 33 (61.1) | 13 (56.5) | 46 (54.8) | 53 (57.0) | 52 (44.1) | 15 (51.7) |
| | >2 | | 282 (45.5) | 21 (41.2) | 36 (37.5) | 36 (50.0) | 21 (38.9) | 10 (43.5) | 38 (45.2) | 40 (43.0) | 66 (55.9) | 14 (48.3) |
| **LDH, n (%)** | Normal | 9 | 381 (62.4) | 33 (64.7) | 59 (63.4) | 43 (60.6) | 31 (58.5) | 17 (73.9) | 55 (65.5) | 54 (58.1) | 72 (63.2) | 17 (58.6) |
| | 1-2x upper limit of normal | | 177 (29.0) | 17 (33.3) | 26 (28.0) | 23 (32.4) | 16 (30.2) | 3 (13.0) | 21 (25.0) | 33 (35.5) | 31 (27.2) | 7 (24.1) |
| | >2x upper limit of normal | | 53 (8.7) | 1 (2.0) | 8 (8.6) | 5 (7.0) | 6 (11.3) | 3 (13.0) | 8 (9.5) | 6 (6.5) | 11 (9.6) | 5 (17.2) |
| **Durable clinical benefit, n (%)** | No benefit | 0 | 253 (40.8) | 18 (35.3) | 39 (40.6) | 35 (48.6) | 23 (42.6) | 9 (39.1) | 33 (39.3) | 38 (40.9) | 46 (39.0) | 12 (41.4) |
| | Benefit | | 367 (59.2) | 33 (64.7) | 57 (59.4) | 37 (51.4) | 31 (57.4) | 14 (60.9) | 51 (60.7) | 55 (59.1) | 72 (61.0) | 17 (58.6) |
| **Response, n (%)** | No response | 0 | 302 (48.7) | 22 (43.1) | 49 (51.0) | 38 (52.8) | 25 (46.3) | 10 (43.5) | 39 (46.4) | 50 (53.8) | 54 (45.8) | 15 (51.7) |
| | Response | | 318 (51.3) | 29 (56.9) | 47 (49.0) | 34 (47.2) | 29 (53.7) | 13 (56.5) | 45 (53.6) | 43 (46.2) | 64 (54.2) | 14 (48.3) |
| **Therapy, n (%)** | Anti-PD1 | 0 | 370 (59.7) | 30 (58.8) | 49 (51.0) | 55 (76.4) | 37 (68.5) | 15 (65.2) | 50 (59.5) | 51 (54.8) | 64 (54.2) | 19 (65.5) |
| | Ipilimumab & Nivolumab | | 250 (40.3) | 21 (41.2) | 47 (49.0) | 17 (23.6) | 17 (31.5) | 8 (34.8) | 34 (40.5) | 42 (45.2) | 54 (45.8) | 10 (34.5) |

Abbreviations – LUMC: Leids Universitair Medisch Centrum; MST: Medisch Spectrum Twente; UMCU: Universitair Medisch Centrum Utrecht; AUMC: Amsterdam Universitair Medisch Centrum; ECOG: Eastern Cooperative Oncology Group; LDH: lactate dehydrogenase.

#### Supplementary Table 4 - Patient characteristics per treatment subgroup

| | | Missing | Overall | Anti-PD1 | Ipilimumab & Nivolumab |
|---|---|---|---|---|---|
| **n** | | | 620 | 370 | 250 |
| **Age, median [Q1,Q3]** | | 0 | 67.5 [58.0,75.0] | 69.5 [61.0,77.0] | 63.0 [53.0,72.0] |
| **Sex, n (%)** | **Female** | 0 | 239 (38.5) | 146 (39.5) | 93 (37.2) |
| | **Male** | | 381 (61.5) | 224 (60.5) | 157 (62.8) |
| **Stage, n (%)** | **IIIC** | 4 | 25 (4.1) | 22 (6.0) | 3 (1.2) |
| | **M1a** | | 49 (8.0) | 45 (12.2) | 4 (1.6) |
| | **M1b** | | 94 (15.3) | 79 (21.5) | 15 (6.0) |
| | **M1c** | | 296 (48.1) | 170 (46.2) | 126 (50.8) |
| | **M1d** | | 152 (24.7) | 52 (14.1) | 100 (40.3) |
| **ECOG performance status, n (%)** | **0** | 26 | 287 (48.3) | 187 (53.1) | 100 (41.3) |
| | **1** | | 247 (41.6) | 139 (39.5) | 108 (44.6) |
| | **2-4** | | 60 (10.1) | 26 (7.4) | 34 (14.0) |
| **Primary tumor location, n (%)** | **Acral** | 10 | 15 (2.5) | 9 (2.5) | 6 (2.5) |
| | **Extremity** | | 167 (27.4) | 110 (30.0) | 57 (23.5) |
| | **Head, neck** | | 66 (10.8) | 47 (12.8) | 19 (7.8) |
| | **Trunk** | | 247 (40.5) | 139 (37.9) | 108 (44.4) |
| | **Unknown** | | 115 (18.9) | 62 (16.9) | 53 (21.8) |
| **Brain metastases, n (%)** | **Absent** | 45 | 423 (73.6) | 286 (84.6) | 137 (57.8) |
| | **Asymptomatic** | | 76 (13.2) | 27 (8.0) | 49 (20.7) |
| | **Symptomatic** | | 76 (13.2) | 25 (7.4) | 51 (21.5) |
| **Liver metastases, n (%)** | **Absent** | 30 | 398 (67.5) | 266 (77.1) | 132 (53.9) |
| | **Present** | | 192 (32.5) | 79 (22.9) | 113 (46.1) |
| **No. of affected organs, n (%)** | **<3** | 0 | 338 (54.5) | 241 (65.1) | 97 (38.8) |
| | **>2** | | 282 (45.5) | 129 (34.9) | 153 (61.2) |
| **LDH, n (%)** | **Normal** | 9 | 381 (62.4) | 280 (76.7) | 101 (41.1) |
| | **1-2x upper limit of normal** | | 177 (29.0) | 78 (21.4) | 99 (40.2) |
| | **>2x upper limit of normal** | | 53 (8.7) | 7 (1.9) | 46 (18.7) |
| **Durable clinical benefit, n (%)** | **No benefit** | 0 | 253 (40.8) | 143 (38.6) | 110 (44.0) |
| | **Benefit** | | 367 (59.2) | 227 (61.4) | 140 (56.0) |
| **Response, n (%)** | **No response** | 0 | 302 (48.7) | 172 (46.5) | 130 (52.0) |
| | **Response** | | 318 (51.3) | 198 (53.5) | 120 (48.0) |

Abbreviations – ECOG: Eastern Cooperative Oncology Group; LDH: lactate dehydrogenase.

**Supplementary Table 5 - Individual lesion outcomes after 3, 6 and 9 months follow-up**

| | Baseline | 3 months | 6 months | 9 months | Last available followup |
|---|---|---|---|---|---|
| **Total lesions, n (%)** | 2352 (100%) | 1752 (100%) | 1368 (100%) | 1232 (100%) | 1752 (100%) |
| **Has response, n (%)** | | 759 (43.3%) | 914 (66.8%) | 923 (74.9%) | 960 (54.8%) |
| **Has benefit, n (%)** | | 1381 (78.8%) | 1216 (88.8%) | 1155 (93.8%) | 1391 (79.4%) |
| **Missing, n** | | 600 | 984 | 1120 | 600 |

#### Supplementary Table 6 – CT acquisition characteristics per center

| | | Missing | Overall | Amphia | Isala | LUMC | Maxima | MST | Radboud | UMCU | AUMC | Zuyderland |
|---|---|---|---|---|---|---|---|---|---|---|---|---|
| **n** | | | 620 | 51 | 96 | 72 | 54 | 23 | 84 | 93 | 118 | 29 |
| **Current, median [Q1,Q3]** | | 21 | 236 [152,340] | 120 [72,252] | 271 [190,377] | 158 [110,240] | 312 [235,380] | 397 [249,525] | 270 [170,409] | 226 [171,282] | 209 [139,292] | 263 [199,346] |
| **Voltage, median [Q1,Q3]** | | 11 | 120 [100,120] | 100 [100,100] | 120 [120,120] | 120 [120,120] | 100 [100,100] | 100 [100,120] | 100 [100,120] | 120 [120,120] | 120 [100,120] | 120 [120,120] |
| **Slice thickness, median [Q1,Q3]** | | 11 | 1.0 [0.9,3.0] | 3.0 [3.0,3.0] | 1.0 [0.9,1.2] | 1.0 [1.0,1.0] | 3.0 [3.0,3.0] | 3.0 [3.0,3.0] | 1.0 [1.0,1.0] | 0.9 [0.9,1.0] | 1.0 [0.9,2.0] | 3.0 [1.7,5.0] |
| **In-slice pixel spacing, median [Q1,Q3]** | | 10 | 0.8 [0.7,0.8] | 0.8 [0.7,0.9] | 0.8 [0.7,0.9] | 0.8 [0.8,0.9] | 0.5 [0.5,0.7] | 0.8 [0.8,0.8] | 0.8 [0.7,0.9] | 0.8 [0.7,0.8] | 0.8 [0.7,0.9] | 0.8 [0.8,0.9] |
| **Vendor, n (%)** | **Canon Medical Systems** | 37 | 4 (0.7) | | | 4 (5.6) | | | | | | nan (nan) |
| | **GE MEDICAL SYSTEMS** | | 35 (6.0) | 2 (3.9) | 6 (6.7) | 3 (4.2) | 5 (9.3) | 1 (4.3) | 4 (4.8) | | 14 (11.9) | nan (nan) |
| | **Philips** | | 258 (44.3) | 6 (11.8) | 70 (78.7) | 4 (5.6) | 49 (90.7) | | 16 (19.0) | 71 (76.3) | 42 (35.6) | nan (nan) |
| | **Philips Medical Systems** | | 1 (0.2) | 1 (2.0) | | | | | | | | nan (nan) |
| | **SIEMENS** | | 170 (29.2) | 42 (82.4) | 7 (7.9) | 3 (4.2) | | 22 (95.7) | 19 (22.6) | 18 (19.4) | 59 (50.0) | nan (nan) |
| | **TOSHIBA** | | 115 (19.7) | | 6 (6.7) | 57 (80.3) | | | 45 (53.6) | 4 (4.3) | 3 (2.5) | nan (nan) |
| **Model, n (%)** | **Aquilion** | 37 | 32 (5.5) | | | 20 (28.2) | | | 8 (9.5) | 2 (2.2) | 2 (1.7) | nan (nan) |
| | **Aquilion ONE** | | 61 (10.5) | | 1 (1.1) | 40 (56.3) | | | 19 (22.6) | | 1 (0.8) | nan (nan) |
| | **Aquilion PRIME** | | 8 (1.4) | | 5 (5.6) | 1 (1.4) | | | | 2 (2.2) | | nan (nan) |
| | **Aquilion Precision** | | 18 (3.1) | | | | | | 18 (21.4) | | | nan (nan) |
| | **Biograph 16** | | 9 (1.5) | | | | | | | | 9 (7.6) | nan (nan) |
| | **Biograph 40** | | 4 (0.7) | | | | | 2 (8.7) | 1 (1.2) | | 1 (0.8) | nan (nan) |
| | **Biograph128** | | 2 (0.3) | 1 (2.0) | | | | | | | 1 (0.8) | nan (nan) |
| | **Biograph128 Edge** | | 3 (0.5) | | | | | | | | 3 (2.5) | nan (nan) |
| | **Biograph40** | | 12 (2.1) | 1 (2.0) | 2 (2.2) | | | | 5 (6.0) | 3 (3.2) | 1 (0.8) | nan (nan) |
| | **Biograph64** | | 33 (5.7) | 22 (43.1) | | | | | | | 11 (9.3) | nan (nan) |
| | **Brilliance 16** | | 1 (0.2) | | 1 (1.1) | | | | | | | nan (nan) |

| | | | | | | | | | | | | |
|---|---|---|---|---|---|---|---|---|---|---|---|---|
| | **Brilliance 40** | | 2 (0.3) | | | | | | 2 (2.4) | | | nan (nan) |
| | **Brilliance 64** | | 28 (4.8) | | | | | | 4 (4.8) | 22 (23.7) | 2 (1.7) | nan (nan) |
| | **Discovery 710** | | 5 (0.9) | | | | 5 (9.3) | | | | | nan (nan) |
| | **Discovery CT750 HD** | | 15 (2.6) | | | | | | 1 (1.2) | | 14 (11.9) | nan (nan) |
| | **Discovery MI** | | 2 (0.3) | | | 1 (1.4) | | 1 (4.3) | | | | nan (nan) |
| | **Discovery STE** | | 4 (0.7) | | 4 (4.5) | | | | | | | nan (nan) |
| | **Emotion 6** | | 1 (0.2) | | | | | | | 1 (1.1) | | nan (nan) |
| | **GEMINI TF TOF 16** | | 1 (0.2) | 1 (2.0) | | | | | | | | nan (nan) |
| | **GEMINI TF TOF 64** | | 2 (0.3) | | | | | | 1 (1.2) | | 1 (0.8) | nan (nan) |
| | **GEMINI TF TOF 64T** | | 1 (0.2) | | | 1 (1.4) | | | | | | nan (nan) |
| | **IQon - Spectral CT** | | 19 (3.3) | | | | | | | 19 (20.4) | | nan (nan) |
| | **Ingenuity CT** | | 46 (7.9) | 1 (2.0) | 16 (18.0) | | 28 (51.9) | | 1 (1.2) | | | nan (nan) |
| | **Ingenuity Core** | | 2 (0.3) | | | 2 (2.8) | | | | | | nan (nan) |
| | **Ingenuity Core 128** | | 2 (0.3) | | | | | | | | 2 (1.7) | nan (nan) |
| | **Ingenuity TF PET/CT** | | 23 (3.9) | | 23 (25.8) | | | | | | | nan (nan) |
| | **Ingenuity TF PET/CT** | | 24 (4.1) | 2 (3.9) | 7 (7.9) | | | | | | 15 (12.7) | nan (nan) |
| | **LightSpeed VCT** | | 4 (0.7) | 2 (3.9) | | 2 (2.8) | | | | | | nan (nan) |
| | **Mx8000 IDT 16** | | 1 (0.2) | | | | 1 (1.9) | | | | | nan (nan) |
| | **Optima CT660** | | 5 (0.9) | | 2 (2.2) | | | | 3 (3.6) | | | nan (nan) |
| | **SOMATOM Definition AS** | | 33 (5.7) | 14 (27.5) | 1 (1.1) | | | 4 (17.4) | 4 (4.8) | 6 (6.5) | 4 (3.4) | nan (nan) |
| | **SOMATOM Definition AS+** | | 3 (0.5) | | | | | 2 (8.7) | | | 1 (0.8) | nan (nan) |
| | **SOMATOM Definition Edge** | | 4 (0.7) | | 3 (3.4) | | | | 1 (1.2) | | | nan (nan) |
| | **SOMATOM Definition Flash** | | 27 (4.6) | 2 (3.9) | 1 (1.1) | 2 (2.8) | | 14 (60.9) | 4 (4.8) | 3 (3.2) | 1 (0.8) | nan (nan) |
| | **SOMATOM Drive** | | 6 (1.0) | | | | | | | | 6 (5.1) | nan (nan) |

| | | | | | | | | | | | | |
|---|---|---|---|---|---|---|---|---|---|---|---|---|
| | **SOMATOM Edge Plus** | | 3 (0.5) | 2 (3.9) | | 1 (1.4) | | | | | | nan (nan) |
| | **SOMATOM Force** | | 15 (2.6) | | | | | | | 5 (5.4) | 10 (8.5) | nan (nan) |
| | **Sensation 64** | | 14 (2.4) | | | | | | 3 (3.6) | | 11 (9.3) | nan (nan) |
| | **TruFlight Select** | | 1 (0.2) | 1 (2.0) | | | | | | | | nan (nan) |
| | **Vereos PET/CT** | | 12 (2.1) | | 8 (9.0) | | | | | | 4 (3.4) | nan (nan) |
| | **iCT 256** | | 94 (16.1) | 2 (3.9) | 15 (16.9) | 1 (1.4) | 20 (37.0) | | 8 (9.5) | 30 (32.3) | 18 (15.3) | nan (nan) |
| | **syngo.via.VB 20A** | | 1 (0.2) | | | | | | 1 (1.2) | | | nan (nan) |

Abbreviations – LUMC: Leids Universitair Medisch Centrum; MST: Medisch Spectrum Twente; UMCU: Universitair Medisch Centrum Utrecht; AUMC: Amsterdam Universitair Medisch Centrum.

#### Supplementary Table 7 – Selected models per fold

| | Model | Preprocessor | Classifier | Hyperparameters | clinical_beta | radiomics_beta | intercept | Best mean AUC in inner loop | Validation AUC |
|---|---|---|---|---|---|---|---|---|---|
| **Amphia** | Radiomics | pca | xgb | ct__correlation_threshold: 0.7158481856169713, lods__level_of_detail: 3, pca__n_components: 13, xgb__learning_rate: 0.36633551871825853, xgb__gamma: 3.1132988947879707, xgb__max_depth: 8, xgb__n_estimators: 84, xgb__reg_alpha: 2.362897797106436, xgb__reg_lambda: 2.594360697766747, aggregation: mean | | | | 0.684 | 0.443 |
| | Clinical | identity | svc | svc__C: 0.13066768311238214, svc__gamma: 0.000516168052739027 | | | | 0.670 | 0.630 |
| | Ensemble | | | | 3.051 | 6.538 | -6.570 | | 0.512 |
| **Isala** | Radiomics | identity | rfc | ct__correlation_threshold: 0.11609293233748338, lods__level_of_detail: 3, rfc__max_features: 7, rfc__max_depth: 4, rfc__n_estimators: 987, aggregation: min | | | | 0.664 | 0.621 |
| | Clinical | pca | svc | pca__n_components: 4, svc__C: 0.3320559103751956, svc__gamma: 0.014077923139972392 | | | | 0.686 | 0.655 |
| | Ensemble | | | | 3.169 | 2.245 | -3.087 | | 0.649 |
| **LUMC** | Radiomics | identity | rfc | ct__correlation_threshold: 0.9794636404890468, lods__level_of_detail: 1, rfc__max_features: 19, rfc__max_depth: 3, rfc__n_estimators: 655, aggregation: min | | | | 0.678 | 0.618 |
| | Clinical | pca | svc | pca__n_components: 3, svc__C: 0.02496474954161821, svc__gamma: 0.015522379296262526 | | | | 0.696 | 0.512 |
| | Ensemble | | | | 3.514 | 2.669 | -3.635 | | 0.528 |
| **Maxima MC** | Radiomics | identity | lr | ct__correlation_threshold: 0.061337225254068733, lods__level_of_detail: 1, lr__alpha: 0.07408236091073953, lr__l1_ratio: 0.21814814885417289, aggregation: min | | | | 0.684 | 0.595 |
| | Clinical | pca | svc | pca__n_components: 4, svc__C: 14.92218339620031, svc__gamma: 0.002863175270831315 | | | | 0.672 | 0.685 |
| | Ensemble | | | | 3.609 | 1.991 | -3.326 | | 0.700 |

| | | | | | | | | | |
|---|---|---|---|---|---|---|---|---|---|
| **MST** | Radiomics | identity | lr | ct__correlation_threshold: 0.10002186634863805, lods__level_of_detail: 1, lr__alpha: 0.05914330202893597, lr__l1_ratio: 0.480588133886884, aggregation: min | | | | 0.632 | 0.810 |
| | Clinical | pca | lr | pca__n_components: 11, lr__alpha: 0.592414568902801, lr__l1_ratio: 0.046450412719997725 | | | | 0.670 | 0.679 |
| | Ensemble | | | | 7.420 | 5.351 | -8.285 | | 0.833 |
| **Radboud** | Radiomics | identity | rfc | ct__correlation_threshold: 0.544261038412369, lods__level_of_detail: 3, rfc__max_features: 10, rfc__max_depth: 6, rfc__n_estimators: 821, aggregation: min | | | | 0.657 | 0.567 |
| | Clinical | identity | lr | lr__alpha: 0.007547319415538378, lr__l1_ratio: 0.011004654538390723 | | | | 0.701 | 0.499 |
| | Ensemble | | | | 3.343 | 1.936 | -2.997 | | 0.519 |
| **UMCU** | Radiomics | pca | xgb | ct__correlation_threshold: 0.13740915907575493, lods__level_of_detail: 3, pca__n_components: 18, xgb__learning_rate: 0.19036017860420434, xgb__gamma: 4.063050667051582, xgb__max_depth: 11, xgb__n_estimators: 36, xgb__reg_alpha: 4.059409467856439, xgb__reg_lambda: 4.729842671189806, aggregation: min | | | | 0.666 | 0.696 |
| | Clinical | pca | svc | pca__n_components: 3, svc__C: 1.4851877511086218, svc__gamma: 0.003509841744352197 | | | | 0.678 | 0.660 |
| | Ensemble | | | | 3.170 | 3.750 | -4.280 | | 0.703 |
| **VUMC** | Radiomics | agglom | xgb | ct__correlation_threshold: 0.909368181361747, lods__level_of_detail: 5, agglom__n_clusters: 5, xgb__learning_rate: 0.6740884752314795, xgb__gamma: 4.40735248739188, xgb__max_depth: 4, xgb__n_estimators: 88, xgb__reg_alpha: 1.0999525824339218, xgb__reg_lambda: 2.54685496820604, aggregation: min | | | | 0.666 | 0.586 |
| | Clinical | pca | svc | pca__n_components: 3, svc__C: 0.2888838362365318, svc__gamma: 0.0015782327810795573 | | | | 0.676 | 0.672 |
| | Ensemble | | | | 2.748 | 4.987 | -4.950 | | 0.648 |

| | | | | | | | | | |
|---|---|---|---|---|---|---|---|---|---|
| **Zuyderland** | Radiomics | pca | rfc | ct__correlation_threshold: 0.47108788319083317, lods__level_of_detail: 3, pca__n_components: 16, rfc__max_features: 12, rfc__max_depth: 4, rfc__n_estimators: 996, aggregation: min | | | | 0.655 | 0.529 |
| | Clinical | pca | svc | pca__n_components: 3, svc__C: 0.8095563021580924, svc__gamma: 0.006590054491061926 | | | | 0.660 | 0.824 |
| | Ensemble | | | | 3.405 | 1.833 | -2.916 | | 0.632 |

Abbreviations – AUC: area under the receiver-operator characteristics curve; LUMC: Leids Universitair Medisch Centrum; MST: Medisch Spectrum Twente; UMCU: Universitair Medisch Centrum Utrecht; AUMC: Amsterdam Universitair Medisch Centrum; lr: logistic regression; pca: principal component analysis; svc: support vector classifier; rfc: random forest classifier; xgb: extreme gradient boosting classifier.

#### Supplementary Table 8 – Cross validated AUROCs for predicting durable clinical benefit in the full dataset using various methods of calculation

| | Radiomics model | | | | | Clinical model | | | | | Combination model | | | | |
|---|---|---|---|---|---|---|---|---|---|---|---|---|---|---|---|
| **Center** | AUC | Equal weight | Patients (N) | Events (N) | Inverse variance | AUC | Equal weight | Patients (N) | Events (N) | Inverse variance | AUC | Equal weight | Patients (N) | Events (N) | Inverse variance |
| Amphia | 0.443 | 1 | 51 | 18 | 138.87 | 0.630 | 1 | 51 | 18 | 126.52 | 0.512 | 1 | 51 | 18 | 122.09 |
| Isala | 0.621 | 1 | 96 | 39 | 302.08 | 0.655 | 1 | 96 | 39 | 306.91 | 0.649 | 1 | 96 | 39 | 289.44 |
| LUMC | 0.618 | 1 | 72 | 35 | 216.45 | 0.512 | 1 | 72 | 35 | 195.83 | 0.528 | 1 | 72 | 35 | 197.87 |
| Maxima | 0.595 | 1 | 54 | 23 | 158.63 | 0.685 | 1 | 54 | 23 | 164.99 | 0.700 | 1 | 54 | 23 | 168.18 |
| MST | 0.810 | 1 | 23 | 9 | 103.74 | 0.679 | 1 | 23 | 9 | 56.18 | 0.833 | 1 | 23 | 9 | 129.91 |
| Radboud | 0.567 | 1 | 84 | 33 | 230.23 | 0.499 | 1 | 84 | 33 | 231.29 | 0.519 | 1 | 84 | 33 | 229.34 |
| UMCU | 0.696 | 1 | 93 | 38 | 342.83 | 0.660 | 1 | 93 | 38 | 298.42 | 0.703 | 1 | 93 | 38 | 325.55 |
| VUMC | 0.586 | 1 | 118 | 46 | 357.44 | 0.672 | 1 | 118 | 46 | 372.51 | 0.648 | 1 | 118 | 46 | 339.54 |
| Zuyderland | 0.529 | 1 | 29 | 12 | 67.35 | 0.824 | 1 | 29 | 12 | 165.89 | 0.632 | 1 | 29 | 12 | 75.93 |
| **AUC using weights** | | **0.607** | **0.604** | **0.606** | **0.613** | | **0.646** | **0.630** | **0.628** | **0.642** | | **0.636** | **0.624** | **0.624** | **0.637** |

Abbreviations – AUC: area under the receiver-operator characteristics curve; LUMC: Leids Universitair Medisch Centrum; MST: Medisch Spectrum Twente; UMCU: Universitair Medisch Centrum Utrecht; AUMC: Amsterdam Universitair Medisch Centrum.

**Supplementary Figure 1 - Random effects pooled analysis of difference in AUROC between ensemble model and clinical model for predicting durable clinical benefit in full dataset**

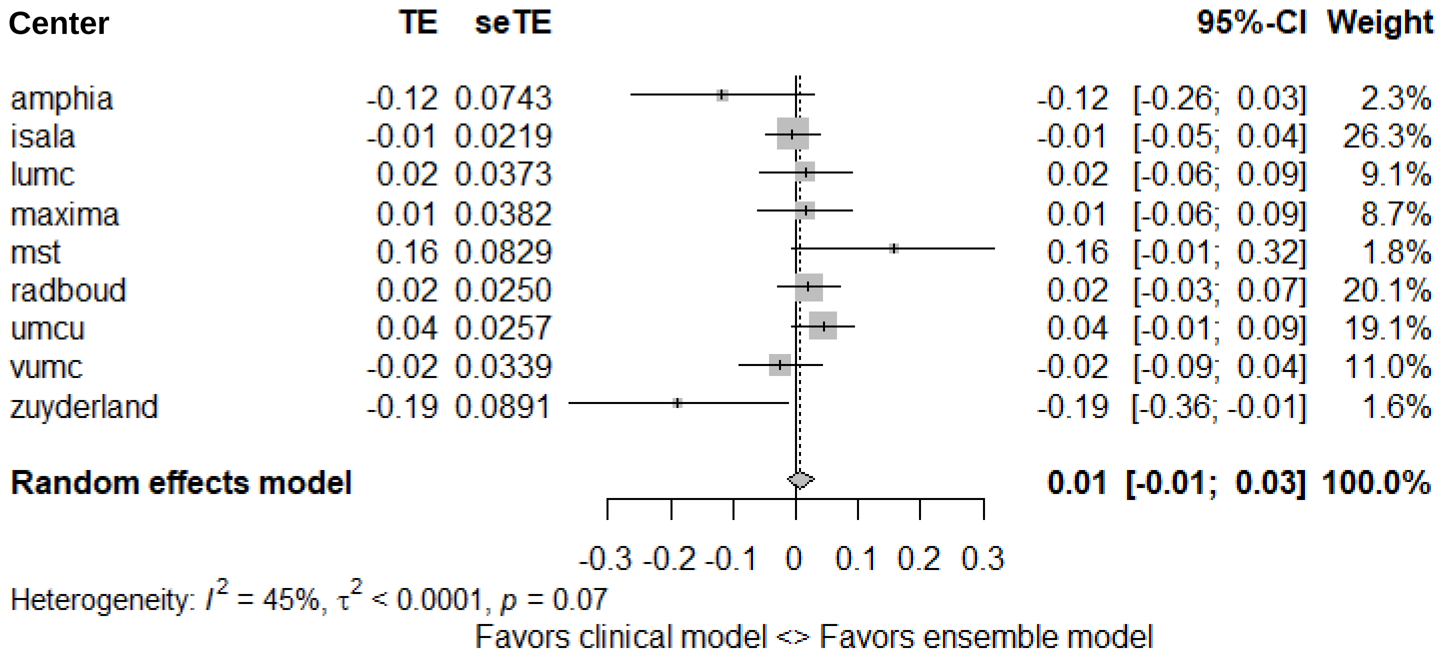

**Supplementary Figure 2 - Random effects pooled analysis of difference in AUROC between ensemble model and clinical model for predicting objective response in full dataset**

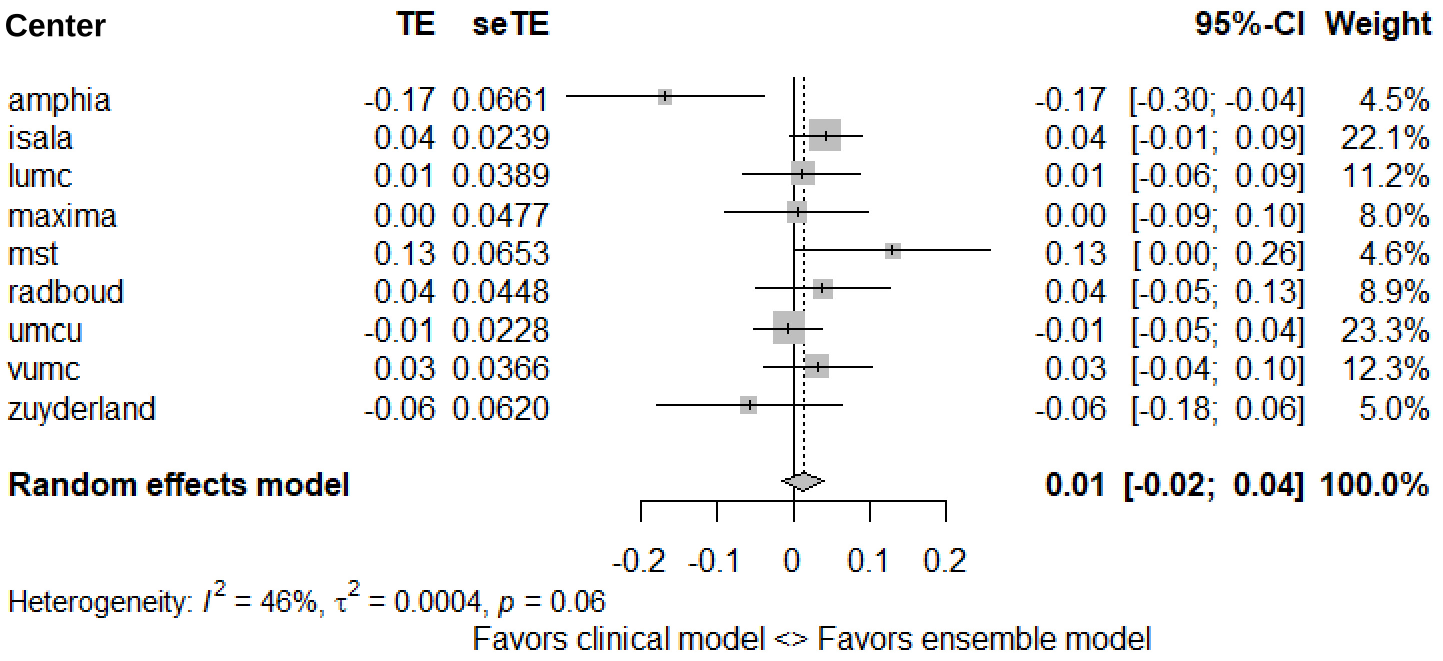

**Supplementary Figure 3 - ROC curves for predicting objective response in all included patients**

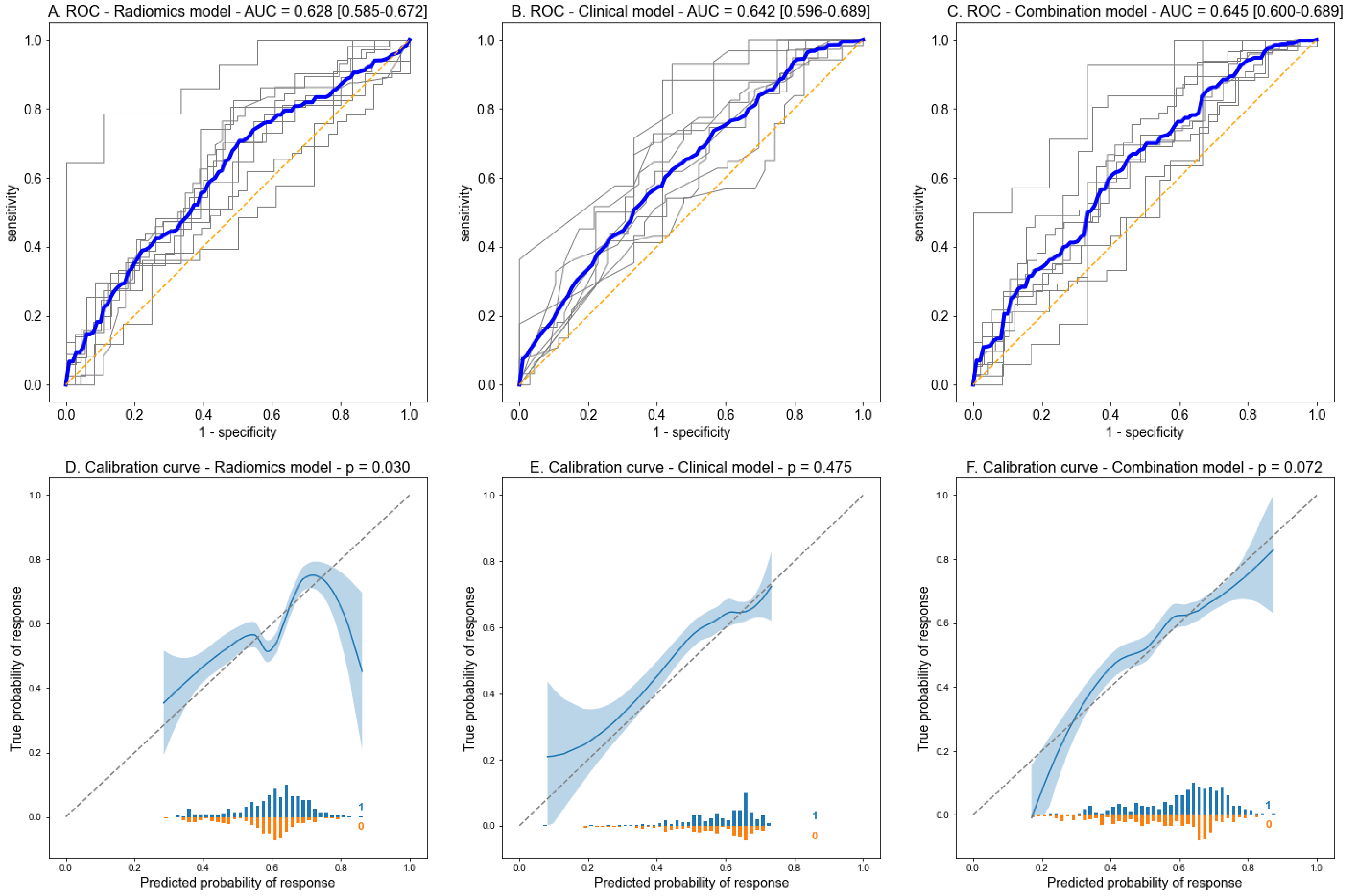

**Supplementary Figure 4 - ROC curves for predicting objective response in subgroup for patients treated with anti-PD1 + anti-CTLA4 therapy**

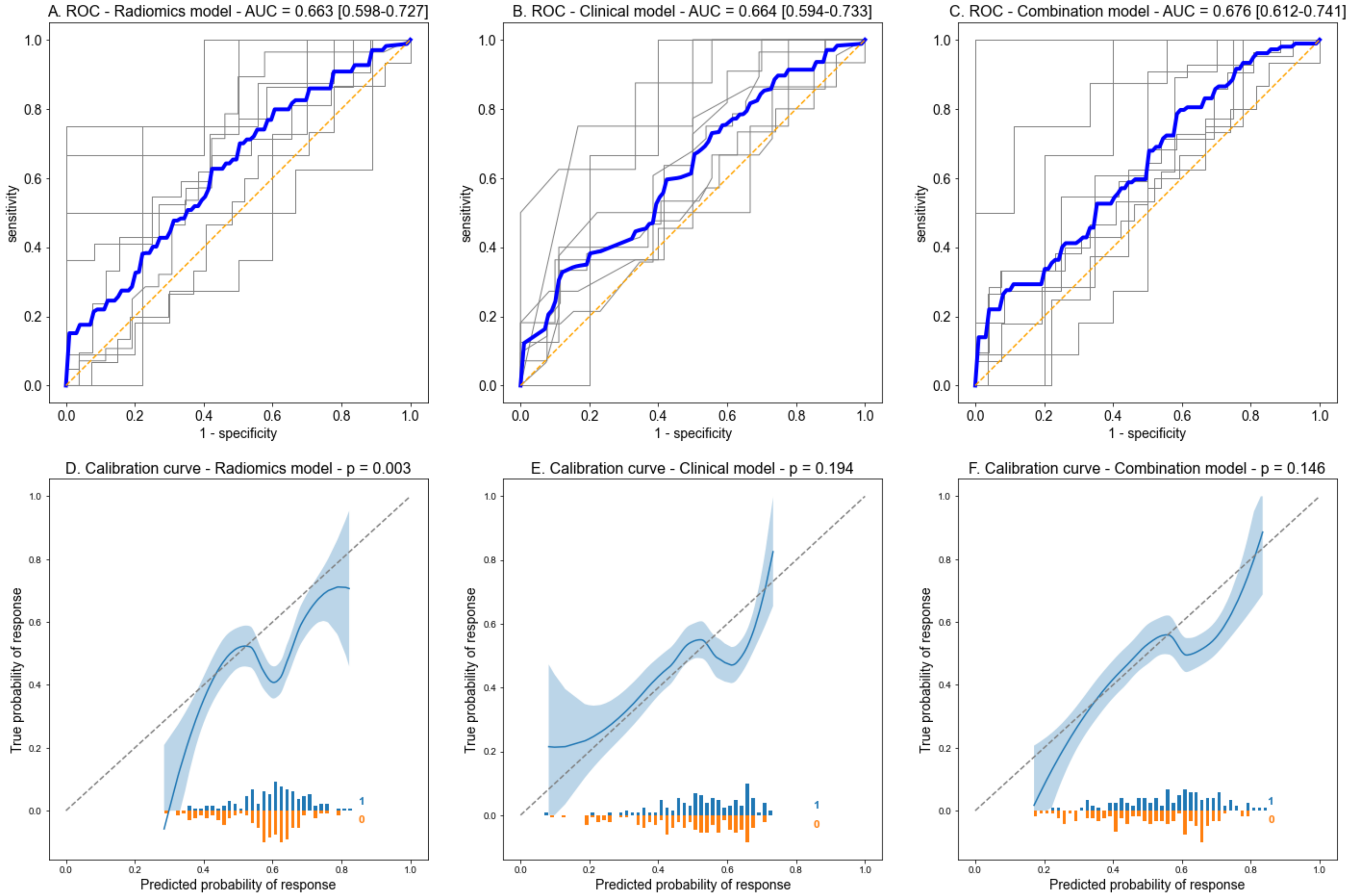

**Supplementary Figure 5 - ROC curves for predicting durable clinical benefit in subgroup for patients treated with anti-PD1 + anti-CTLA4 therapy**

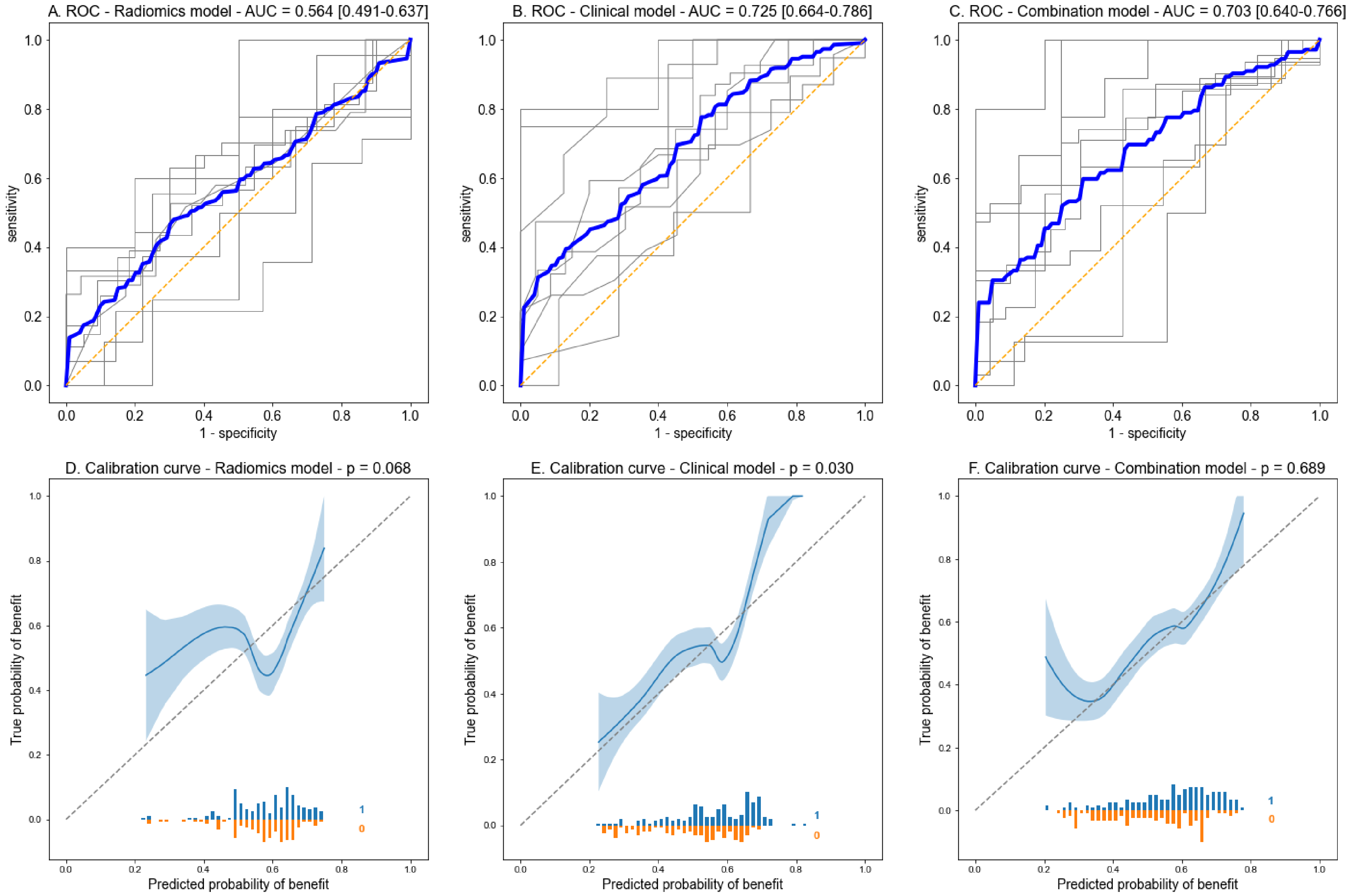

**Supplementary Figure 6 - ROC curves for predicting objective response in subgroup for patients treated with anti-PD1 monotherapy**

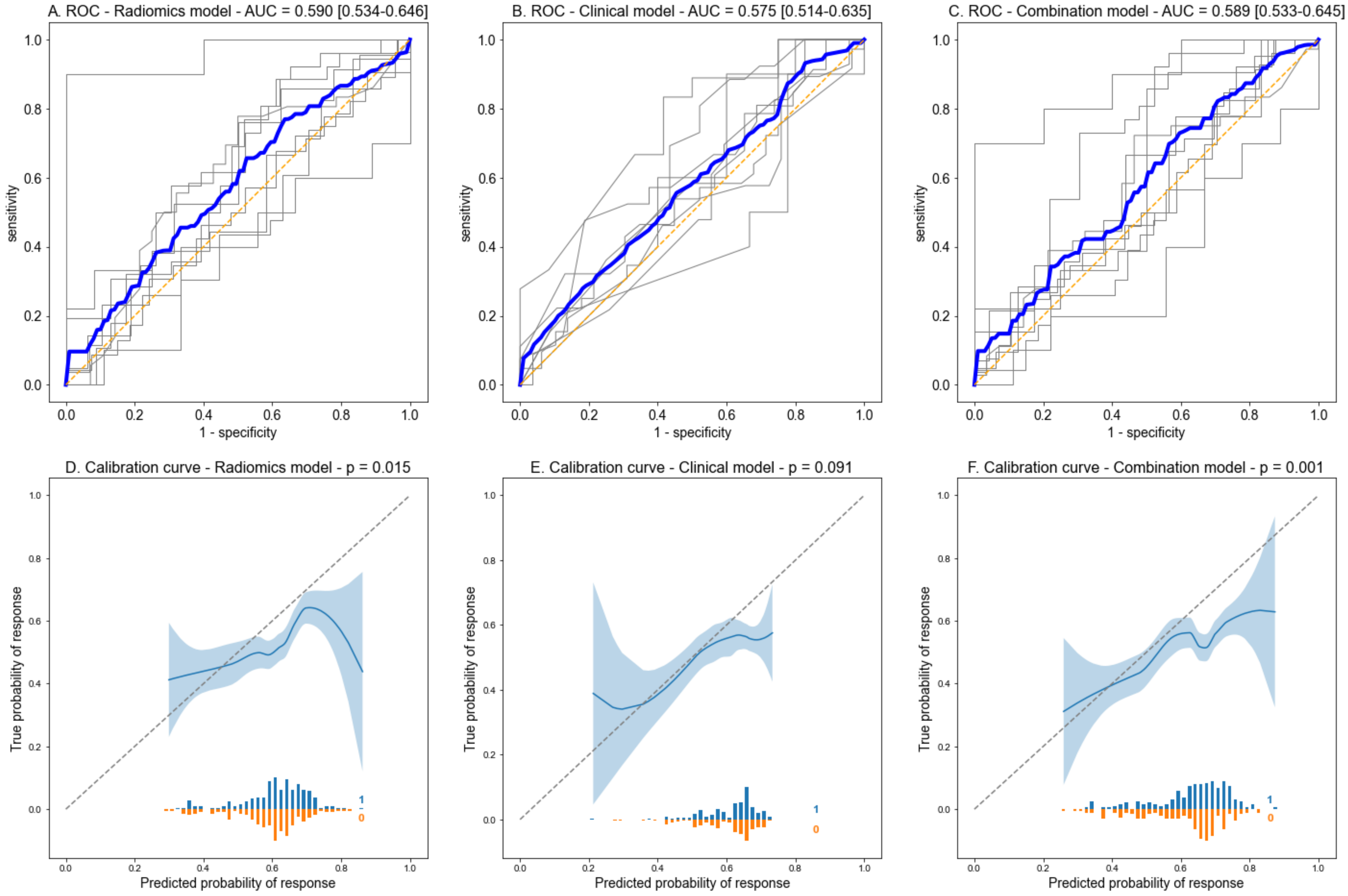

**Supplementary Figure 7 - ROC curves for predicting durable clinical benefit in subgroup for patients treated with anti-PD1 monotherapy**

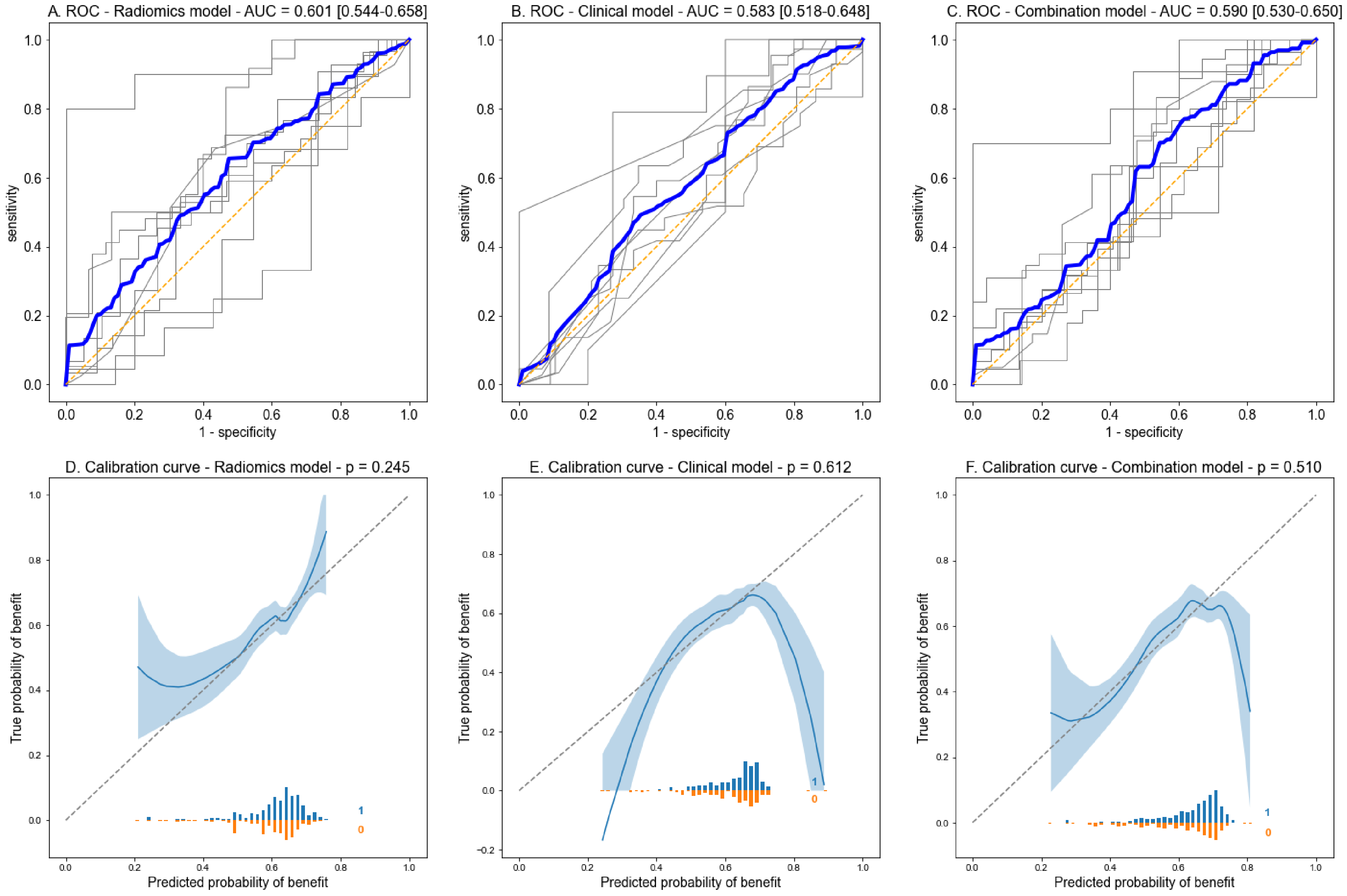
